# High number of SARS-CoV-2 persistent infections uncovered through genetic analysis of samples from a large community-based surveillance study

**DOI:** 10.1101/2023.01.29.23285160

**Authors:** Mahan Ghafari, Matthew Hall, Tanya Golubchik, Daniel Ayoubkhani, Thomas House, George MacIntyre-Cockett, Helen Fryer, Laura Thomson, Anel Nurtay, David Buck, Angie Green, Amy Trebes, Paolo Piazza, Lorne J Lonie, Ruth Studley, Emma Rourke, Darren Smith, Matthew Bashton, Andrew Nelson, Matthew Crown, Clare McCann, Gregory R Young, Rui Andre Nunes dos Santos, Zack Richards, Adnan Tariq, Roberto Cahuantzi, Wellcome Sanger Institute COVID-19 Surveillance Team, COVID-19 Infection Survey Group, The COVID-19 Genomics UK (COG-UK) Consortium, Jeff Barrett, Christophe Fraser, David Bonsall, Ann Sarah Walker, Katrina Lythgoe

**Affiliations:** Big Data Institute, Nuffield Department of Medicine, University of Oxford, Old Road Campus, Oxford OX3 7LF, UK; Department of Biology, University of Oxford, Oxford OX1 3SZ, UK; Sydney Infectious Diseases Institute (Sydney ID), School of Medical Sciences, Faculty of Medicine and Health, University of Sydney; Office for National Statistics, Newport, UK, Medicine and Health, University of Sydney; Leicester Real World Evidence Unit, Diabetes Research Centre, University of Leicester, Leicester, UK; Department of Mathematics, University of Manchester, Manchester, M13 9PL, UK; Wellcome Centre for Human Genetics, Nuffield Department of Medicine, NIHR Biomedical Research Centre, University of Oxford, Old Road Campus, Oxford OX3 7BN, UK; The Hub for Biotechnology in the Built Environment, Department of Applied Sciences, Faculty of Health and Life Sciences, Northumbria University, Newcastle upon Tyne, NE1 8ST, UK; Department of Applied Sciences, Faculty of Health and Life Sciences, Northumbria University, Newcastle upon Tyne, NE1 8ST, UK; Wellcome Sanger Institute, Cambridge CB10 1SA, UK; Nuffield Department of Medicine, University of Oxford, Oxford, UK; The National Institute for Health Research Health Protection Research Unit in Healthcare Associated Infections and Antimicrobial Resistance at the University of Oxford, Oxford, UK; The National Institute for Health Research Oxford Biomedical Research Centre, University of Oxford,Oxford, UK; MRC Clinical Trials Unit at UCL, UCL, London, UK; Oxford University Hospitals NHS Foundation Trust, John Radcliffe Hospital, Headington, Oxford OX3 9DU, UK

## Abstract

Persistent severe acute respiratory syndrome coronavirus 2 (SARS-CoV-2) infections may act as viral reservoirs that could seed future outbreaks ^1–5^, give rise to highly divergent lineages ^6–8^, and contribute to cases with post-acute Coronavirus disease 2019 (COVID-19) sequelae (Long Covid) ^9,10^. However, the population prevalence of persistent infections, their viral load kinetics, and evolutionary dynamics over the course of infections remain largely unknown. We identified 381 infections lasting at least 30 days, of which 54 lasted at least 60 days. These persistently infected individuals had more than 50% higher odds of self-reporting Long Covid compared to the infected controls, and we estimate that 0.09-0.5% of SARS-CoV-2 infections can become persistent and last for at least 60 days. In nearly 70% of the persistent infections we identified, there were long periods during which there were no consensus changes in virus sequences, consistent with prolonged presence of non-replicating virus. Our findings also suggest reinfections with the same major lineage are rare and that many persistent infections are characterised by relapsing viral load dynamics. Furthermore, we found a strong signal for positive selection during persistent infections, with multiple amino acid substitutions in the Spike and ORF1ab genes emerging independently in different individuals, including mutations that are lineage-defining for SARS-CoV-2 variants, at target sites for several monoclonal antibodies, and commonly found in immunocompromised patients ^11–14^. This work has significant implications for understanding and characterising SARS-CoV-2 infection, epidemiology, and evolution.

## Main

The emergence of highly divergent variants of SARS-CoV-2 has been a defining feature of the COVID-19 pandemic. While the evolutionary origins of these variants are still a matter of speculation, multiple pieces of evidence point to chronic persistent infections as their most likely source ^5,7,15^. In particular, infections in immunocompromised patients who cannot clear the virus may lead to persistence for months ^6,7,16,17^ or even years ^8,18^ before potentially seeding new outbreaks in the community ^3^. Persistence of SARS-CoV-2 during chronic infections exposes the viral population to host-immune responses and other selective pressures as a result of treatments over prolonged periods of time. They also release the virus from undergoing the tight population bottlenecks that are characteristic of SARS-CoV-2 transmission ^19,20^, making the viral population less vulnerable to stochastic genetic drift. These adaptive intrahost changes can lead to elevated evolutionary rates, particularly in key regions of the Spike protein that are often associated with immune escape and elevated rates of transmission ^13,14^. Despite the significant public health implications of persistent infections, uncertainty still surrounds how common these infections are among the general population, how long they last, their potential for adaptive evolution, and their contribution to Long Covid.

In this work, we leveraged genetic, symptom, and epidemiological data from the Office for National Statistics Covid Infection Survey (ONS-CIS) ^21^, an ongoing large scale community-based surveillance study carried out in the UK. We identified individuals with persistent SARS-CoV-2 infection and characterised various aspects of their infection including evolutionary changes in the virus, viral load kinetics, number of reported symptoms, and prevalence of Long Covid.

### Identifying persistent infections

We considered more than 100,000 high-quality sequenced samples from the ONS-CIS collected between 2nd November 2020 to 15th August 2022, and representing ∼95,000 people living in ∼75,000 households across the UK (see **Methods**). Individuals in the survey were typically sampled once a week for the first four weeks of their enrolment, and then monthly thereafter. To identify persistent infections we first limited the dataset to individuals with two or more RT-PCR positive samples with cycle threshold (Ct) values ≤30 (in which sequencing was attempted; a proxy for viral load), taken at least 26 days apart, and where the consensus sequences were of the same major lineages of Alpha, Delta, BA.1 or BA.2 (BA.4 and BA.5 not considered). If those sequences also shared the same rare single nucleotide polymorphisms (SNPs) at one or more sites relative to the major-lineage population-level consensus, we classified them as having a persistent infection.

We defined a rare mutation as one observed in 400 or fewer samples within the entire ONS-CIS dataset, giving a false positive rate of identifying persistent infections of 0-3% depending on the major lineage (see **Methods** and **Supplementary Figure 1**). We note that the rare SNP method provides a conservative estimate for the true number of persistent infections since some persistent infections may not have rare mutations. To further evaluate the robustness of our method in identifying persistent infections, we considered the phylogenetic relationship between the sequences from persistent infections relative to other sequences of the same major lineage that belonged to individuals with only a single sequence within the ONS-CIS dataset. The great majority of sets of sequences identified as belonging to the same persistent infection formed monophyletic groups with strong bootstrap support (**Figure 1a;** see also **Supplementary Figure 2**). However, seven sequences did not group with the other sequence(s) from the same persistent infection. All of these had high Ct values (Ct∼30) and poor genome coverage which may explain their lack of clustering on the phylogeny (**Supplementary Figure 2**).

**Figure 1:**
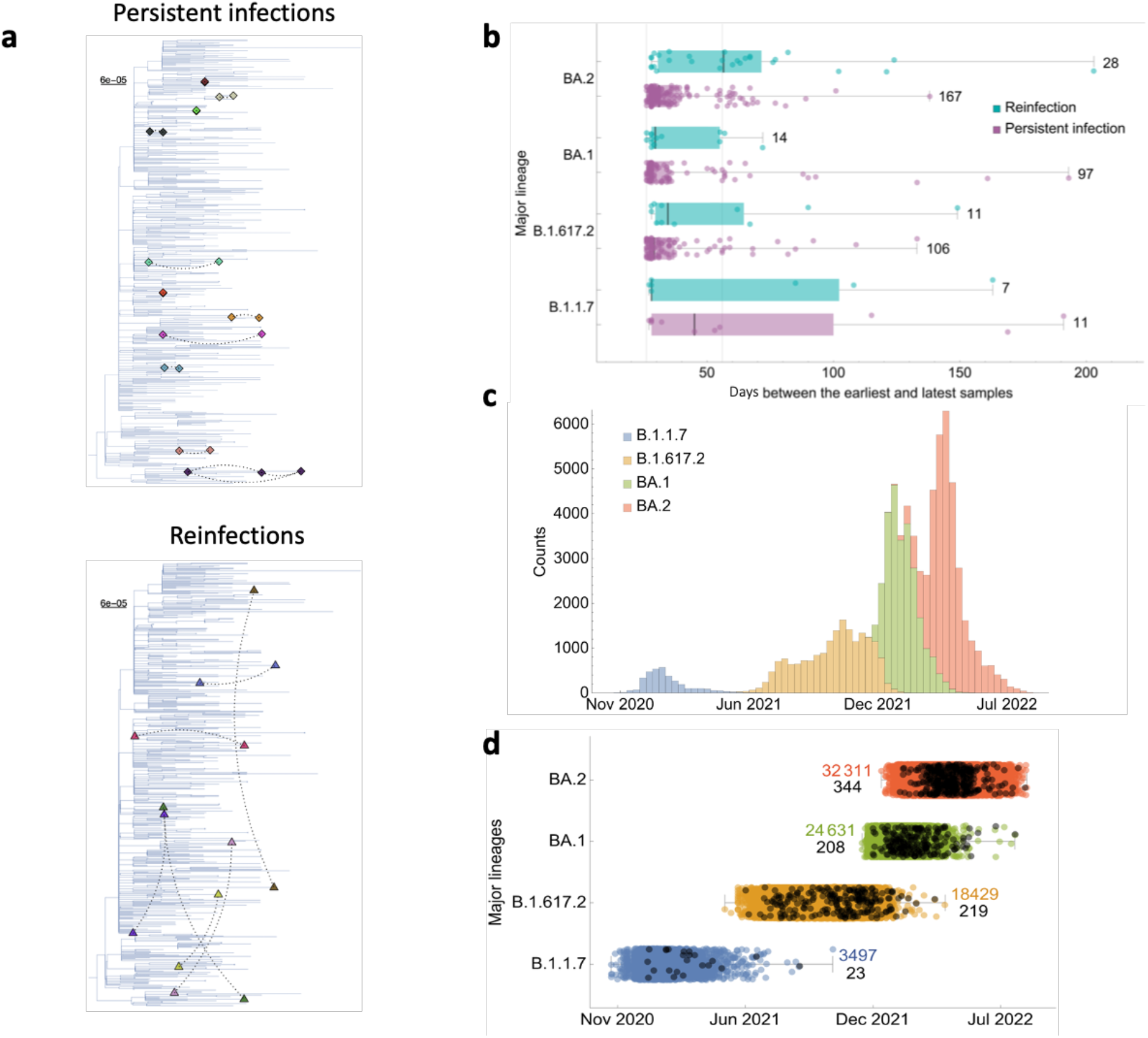
Individuals identified with persistent infections and reinfections within the ONS-CIS. **(a)** Phylogenetic relationship between samples from persistent infections and reinfections with a representative background population of Alpha (see **Supplementary Figure 2** for the analysis on the other three major lineages). Dashed lines connect every pair of sequences from the same individual. Pairs from persistent infections cluster closely together while reinfections do not. All sequences from the same individual are given the same colour. **(b)** Days between the earliest and latest genomic samples from persistent infections and reinfections. Each point represents a single individual. Solid vertical lines show the 26- and 56-day cutoffs. Numbers on the side of each box shows the total counts per category for each major lineage. **(c)** Total number of sequences in the ONS-CIS per major lineage over time. **(d)** Timing of persistent infections (black) during the UK epidemic. Some persistent infections can be identified up to weeks after the lineage has been replaced at the population level. The numbers on the side of each box shows the total sequence counts for each category.

We found 381 persistent infections with sequences spanning at least 26 days, of which 54 spanned at least 56 days, representing nearly 0.07% (54/77,561) of all individuals with one or more sequences (with Ct≤30) of the four major lineages we investigated in this study (**Figure 1b**; see also **Table 1**). Notably, 9% (2/23), 9% (19/219), and 8% (8/208) of sequences from persistent infections with Alpha, Delta, and BA.1, respectively, were sampled weeks after when the corresponding major lineage has dropped to ≤1% frequency of all the ONS-CIS sequences (**Figure 1c**); the longest infection was with BA.1 and lasted for at least 193 days (see **Figure 1b**).

**Table 1:** Frequency of persistent infections and reinfections per major lineage.

| Major lineage | Reinfection >26 days | Reinfection >56 days | Persistent infection >26 days | Persistent infections >56 days | %Reinfection >26 days | %Reinfection >56 days |
| --- | --- | --- | --- | --- | --- | --- |
| B.1.1.7 | 7 | 3 | 11 | 3 | 39% | 50% |
| B.1.617.2 | 11 | 4 | 106 | 13 | 9% | 24% |
| BA.1 | 14 | 2 | 97 | 15 | 13% | 12% |
| BA.2 | 28 | 15 | 167 | 23 | 14% | 40% |

The actual duration of persistent infections are likely to be at least 3-4 days longer than the time between when the first and last sequenced samples were collected, since it typically takes 3-4 days since the start of infection for viral loads to be sufficiently high to be sequenced (Ct≤30) ^22,23^, and similarly viral loads will be too low (Ct values too high) to sequence at the tail end of infection. Since individuals were typically sampled weekly during the first four weeks of enrollment, followed by monthly sampling thereafter, it is unsurprising that most persistent infections had observable durations clustering around 30 or 60 days (see **Supplementary Figure 3**).

We found evidence suggestive of transmission from 11 persistently infected individuals: we identified two households each with two members having concurrent persistent infections, and in nine other households, one member tested positive for SARS-CoV-2 by RT-PCR within 10 days of a positive sequenced sample being collected from a persistently infected individual from the same household. As well as being clustered in households, the sequences from the suspected persistently infected source and the recipient had either no consensus nucleotide differences (nine cases) or one consensus nucleotide difference (two cases), consistent with transmission. If these do represent transmission, it is worth noting that none of these persistent infections involved highly divergent sequences relative to their first sequence, and hence these do not provide an example of the emergence and spread of highly divergent lineages.

### Identifying reinfections with the same major lineage

We considered a pair of sequences from the same individual to indicate a reinfection if they were sampled at least 26 days apart, had at least one consensus nucleotide difference between the sequenced sampling timepoints, and shared no rare SNPs (see **Methods**). This criterion may overestimate the true number of reinfections as some persistent infections may not have a rare SNP, and within-host evolution can lead to the loss of a rare SNP and/or the gain of other mutations leading to differences in the consensus sequence between the samples. We identified three individuals for which pairs of sequences from different sampling timepoints had no identical rare SNPs and at least one consensus difference, but whose viral load trajectories were consistent with a persistent chronic infection. We therefore excluded these individuals from the reinfection group (see **Supplementary Figure 4**).

Overall, we identified 60 reinfections with the same major lineage (**Table 1**). Of all the cases classed as either persistent infections or reinfections with the same major lineage,10-15% were classed as reinfections (see **Table 1**), rising to 20-40% if only samples collected at least 56 days apart were included (see **Figure 1b**). This suggests the number of individuals reinfected with the same major lineage is low compared to the number of individuals with persistent infection. Sequences from individuals identified as reinfected, collected at the point of primary infection and reinfection, did not form monophyletic groups and mostly belonged to distantly related subclades, and hence supports our method for identifying reinfections (**Figure 1a**; **Supplementary Figure 2**).

### Evidence of non-replicating virus during infection

Of the 381 persistently infected individuals that we identified, nearly 70% (267/381) had a pair of sequenced samples taken at least 26 days apart with no nucleotide differences at the consensus level. This is striking given the between-host within-lineage evolutionary rate of SARS-CoV-2 of ∼2 single nucleotide variants (SNVs) per month ^24–26^. We contrasted the number of consensus nucleotide differences between pairs of samples from persistent infections with 16,000 random pairs of sequences sampled from the complete set of sequenced samples from the ONS-CIS, and with each pair from the same major lineage (**Supplementary Figure 5**). Of these, only 6 pairs had no SNVs (i.e., less than 0.04% of pairs).

### Emergence of notable mutations

For all pairs of sequences from each of the persistent infections, we identified mutations for which there was a change in consensus between the two sampling time points. Among the 381 persistently infected individuals, we observed 317 changes in the consensus nucleotide representing 277 unique mutations, and 31 deletions representing 18 unique deletions. Many of these mutations have previously been identified as either lineage-defining mutations for variants of concern or variants of interest ^27^ (eight mutations and two deletions), recurrent mutations in immunocompromised individuals ^12–14^ (15 mutations and four deletions), or key mutations with antibody escape properties and target sites for various different monoclonal antibodies ^11,28^ (seven mutations) (**Supplementary Table 1**).

We observed several mutations at the same genomic positions in multiple individuals over the course of their persistent infections. For example, three BA.2 infected individuals from different households acquired a mutation at codon position 547 in Spike (**Figure 2**), two of which were the T547K mutation which is a lineage-defining mutation for BA.1, and one the K547T mutation (**Figure 2c**; also see **Supplementary Table 1**). Strikingly, twelve individuals acquired a deletion (ORF1ab: Δ81-87) in the NSP1 coding region. A similar deletion has previously been observed during the chronic infection of an immunocompromised individual with cancer ^16^, and was associated with lower type I interferon response in infected cells ^29^. We also identified a persistent infection with BA.1 lasting for at least 133 days during which 33 unique mutations (23 mutations in ORF1ab, 6 in Spike, 1 in ORF3a, 1 in M, and 2 in ORF7) were observed (see **Supplementary Figure 2**); eleven of the ORF1ab mutations and all of the mutations in Spike, ORF3a, and ORF7 were nonsynonymous.

**Figure 2:**
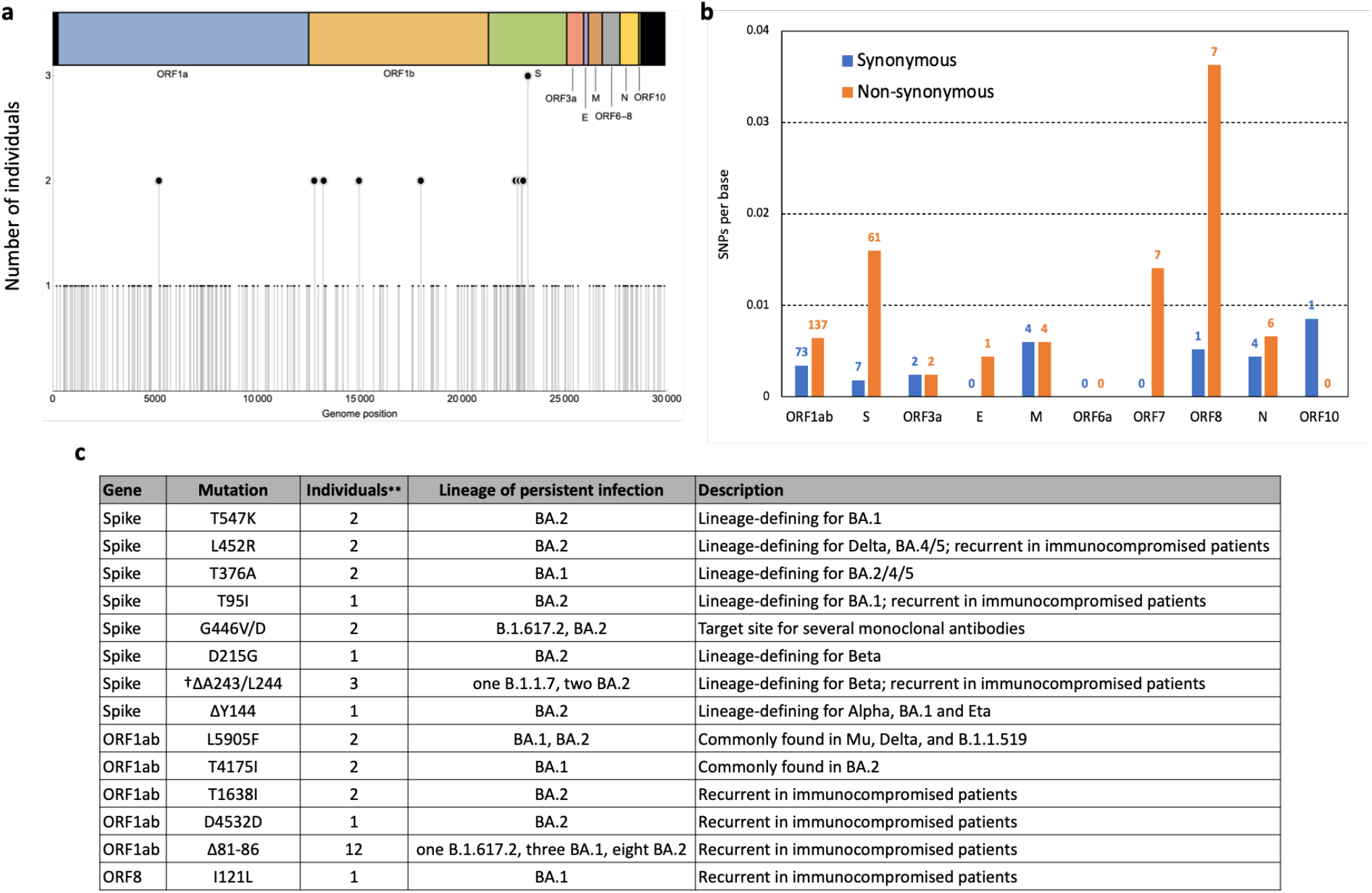
Distribution of single nucleotide polymorphisms and non-synonymous vs. synonymous mutations detected during persistent infections. **(a)** Frequency of mutations that resulted in a SNP change during one or more persistent infections. **(b)** Number of synonymous (blue) and non-synonymous (orange) mutations per gene during persistent infections. Numbers on each column show the total counts of SNPs in each category of mutations. **(c)** Description of recurrent mutations and deletions identified during persistent infections. See **Supplementary Table 1** for information about other mutations. **None of the recurrent mutations were from households with other infections. †Δ represents a deletion

Overall, we observed a strong signal for positive selection in Spike, with nearly nine-fold more nonsynonymous compared to synonymous mutations (**Figure 2b**). With a total of seven nonsynonymous mutations, ORF8 had the highest per base number of nonsynonymous mutations followed by Spike with 61 nonsynonymous mutations.

### Persistence with relapsing viral load

Of the 381 persistent infections, 65 had three or more RT-PCR tests taken over the course of their infection. We classified these infections as persistent-relapsing if they had a negative RT-PCR test during the infection (n=20), and the rest as persistent-chronic (n=45) (**Figure 3a,b**). Given the weekly or monthly sampling of individuals enrolled in the ONS-CIS, infections classed as persistent-chronic may have unsampled periods of very low viral burden, meaning the persistent-relapsing category is likely to be an underestimate. Nonetheless, the observation of relapsing viral load dynamics in over 30% of cases is striking given that, in the absence of genetic information, they could have been misidentified as reinfections, depending on the definition used. Of the 27 cases identified as reinfections with three or more RT-PCR tests, all showed relapsing viral load dynamics (**Figure 3c**).

**Figure 3:**
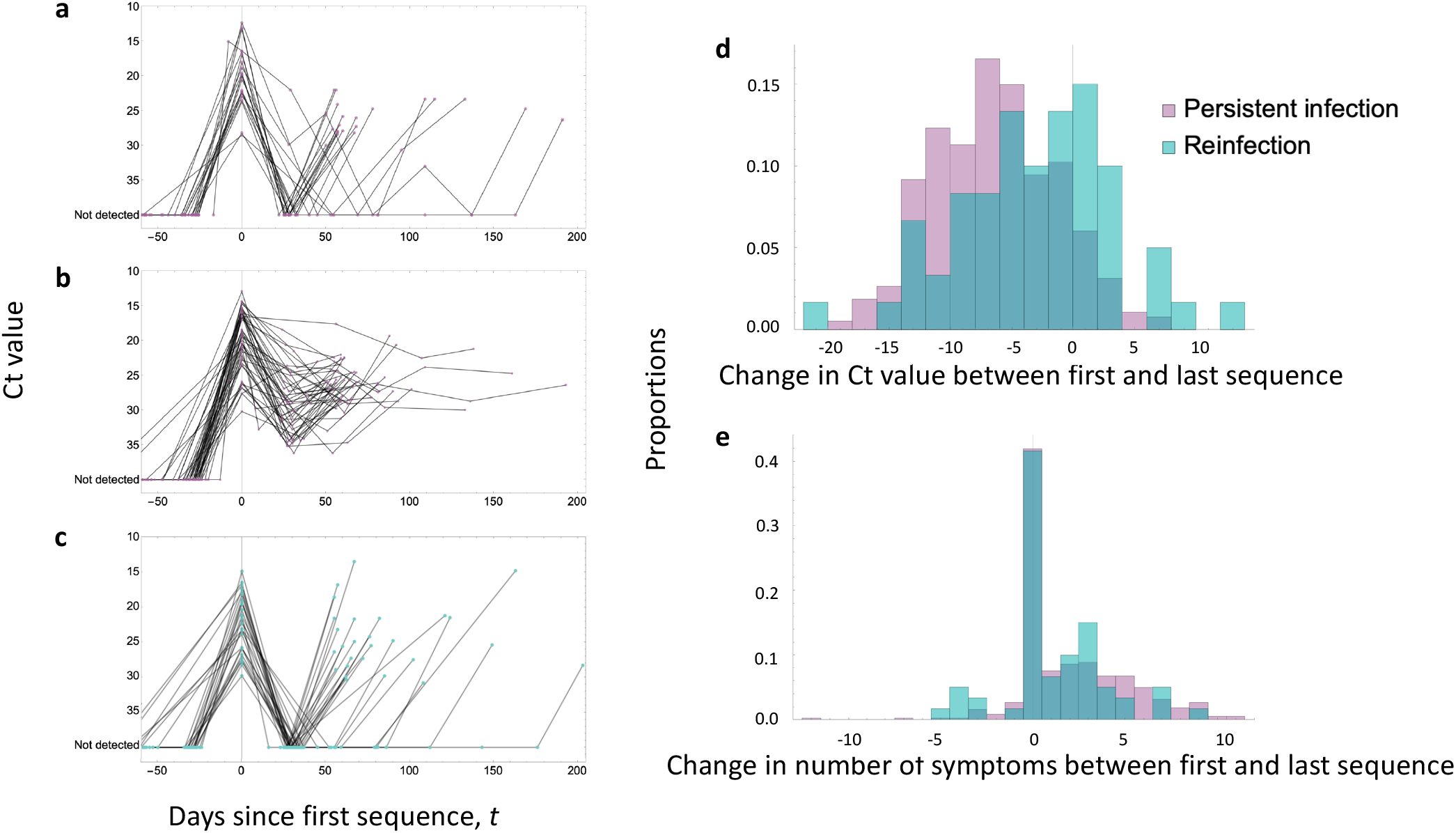
Comparison of viral load dynamics and number of reported symptoms during persistent infections and reinfections. Viral load trajectories of persistent infections with **(a)** relapsing and **(b)** chronic persistent infections and **(c)** reinfections with at least three PCR tests taken over the course of infection/until reinfection. **(d)** Change in cycle threshold (Ct) value and **(e)** total number of symptoms reported between the first and last time points with sequenced samples for all 381 persistent infections (purple) and 60 reinfections (cyan). Viral load at the last time point is significantly lower for persistent infections with Ct value being more than +6.7 (IQR: +3.2,+10.2) units higher at the last time point (paired t-test p<10^−8^). The difference is less pronounced for reinfections with +2.5 (IQR: -1.1, +7.4) units difference between primary infection and reinfections (paired t-test p=0.0003). Similarly, persistently infected individuals tend to report fewer number of symptoms at the later stages of their infection compared to reinfections in the last 7 days at the last time point (at which a sequence was obtained) when compared to the first. On average, persistently infected individuals report two more symptoms at the time of their first sequenceable sample (virus samples with a positive Ct≤30) relative to their last sample, with a median of 1 (IQR: 0, 4) fewer reported symptoms (paired Wilcoxon p<10^−29^), while reinfected individuals report only one more symptom at the point of primary infection relative to reinfection, with a median of 0 (IQR: 0, 3) fewer reported symptoms (paired Wilcoxon p=0.005).

As the sampling strategy of ONS-CIS is based on testing representative individuals across the UK regardless of symptoms, we can estimate the percentage of SARS-CoV-2 infections that are persistent and last for longer than 60 days in the general population. This requires making assumptions about how many persistent infections are missed among ONS-CIS participants due to the monthly (and weekly) sampling. More precisely, estimating the proportion of infections that are persistent depends on the proportion of days the infection has sequenceable virus during the infection (would have Ct≤30 if tested); the fewer the number of days the infection has sequenceable virus, the more likely it is that a persistent infection is be missed. By taking two extreme scenarios for the proportion of days that the virus is sequenceable during persistent infection (0.7 and 0.14; see **Methods**), we estimate that approximately 0.1-0.5% of infections become persistent for 60 days or more.

### Difference in viral load and symptoms

For the majority of persistent infections, Ct values (inversely proportional to viral titre ^30^) were higher at the last sequenced time point compared to the first sequenced time point (**Figure 3d**). For reinfections with the same major lineage, the last sequenced sample also had higher Ct values than the first, but the magnitude of the difference was smaller compared to persistent infections (**Figure 2d**). In both cases, the rise in Ct value (decrease in viral titre) during infections or between reinfections could be a consequence of host immunity or within-host compartmentalisation. Additionally, the rise in Ct for reinfections could be due to the disproportionate sampling of individuals with older infections, which tend to have lower viral loads, towards the end of an epidemic wave ^31,32^.

Individuals with persistent infections remained largely asymptomatic during the later stages of infection, with, on average, reporting two fewer symptoms in the last 7 days at the last time of sampling (at which a sequence was obtained) when compared to the first. They also consistently reported very few or no symptoms after the first positive sample (**Figure 3e**). In comparison, individuals reinfected with the same major lineage reported on average only one fewer symptom at the reinfection sampling time point compared to the primary sampling time point (**Figure 3e**).

### Prevalence of Long Covid

From February 2021, as well as reporting symptoms, participants were asked if they describe themselves as having Long Covid and still experiencing symptoms more than four weeks after they first had COVID-19 (see **Methods**). We estimated the prevalence of self-reported Long Covid in the persistently infected individuals compared with a control group, accounting for several confounding variables (see **Methods**). In the persistent infection group, 9.0% of respondents (32/354) self-reported Long Covid at their first visit ≥12 weeks since the start of infection, and 7.1% (19/266) reported Long Covid at ≥26 weeks. However, in the control group, only 5.1% (4,976/97,404) reported Long Covid at their first visit ≥12 weeks, and 4.5% (3,261/72,407) reported Long Covid at ≥26 weeks.

Correcting for confounders, we found strong evidence for a 55% higher odds of reporting Long Covid ≥12 weeks post-infection among persistently infected individuals compared to controls (Chi square test with Yates correction ^33^ p=0.004 for the unadjusted model, p=0.021 for the adjusted model), but no evidence of a difference for Long Covid ≥26 weeks post-infection (p=0.127 for the unadjusted model, p=0.367 for the adjusted model) (**Table 2**). The lower rate of reporting Long Covid 26 weeks post-infection could be because the majority of the persistent infections we identified lasted for less than 26 weeks, and hence persistence of an infection may no longer be a contributing factor to Long Covid.

**Table 2:** Prevalence of Long Covid in persistently infected individuals.

Long Covid ≥12 weeks post-infection
| Group | *n* | Long Covid | Median follow-up (IQR*) | Unadjusted OR† (95% CI**) | Adjusted OR (95% CI) |
| --- | --- | --- | --- | --- | --- |
| Exposed | 356 | 32 (9.0%) | 101 (91-113) | 1.27 (1.19-2.47) | 1.55 (1.07-2.25) |
| Control | 78,902 | 4,291 (5.4%) | 100 (91-115) | Reference | Reference |

Long Covid ≥26 weeks post-infection
| Group | *n* | Long Covid | Median follow-up (IQR) | Unadjusted OR (95% CI) | Adjusted OR (95% CI) |
| --- | --- | --- | --- | --- | --- |
| Exposed | 326 | 19 (5.8%) | 312 (271-390) | 1.44 (0.90-2.29) | 1.24 (0.77-2.00) |
| Control | 72,608 | 3,000 (4.1%) | 320 (272-384) | Reference | Reference |

\* IQR=interquartile range
†OR=odds ratio
\*\*CI=confidence interval

## Discussion

We developed a robust approach for identifying persistent SARS-CoV-2 infections in individuals with sequenced samples spanning a month or longer. Of the 381 persistent infections we identified among participants of the ONS-CIS, 54 lasted at least two months, two over six months, and in some cases the infecting lineage had gone extinct in the general population. In contrast, we only identified 60 reinfections by the same major lineage as the primary infection, suggesting immunity to the same variant remains strong after infection, at least until the lineage has gone extinct.

The large number of persistent infections we uncovered is striking, given the leading hypothesis that many of the variants of concern (VOCs) emerged wholly or partially during long-term chronic infections in immunocompromised individuals ^1^. Since the ONS-CIS is a community-based surveillance study, our observations suggest the pool of people in which long-term infections could occur, and hence potential sources of divergent variants, may be much larger than generally thought. We estimate that more than nearly one in a thousand of all infections, and potentially as many as one in 200, may become persistent for at least two months. The harbouring of these persistent infections in the general community may also help explain the early detection of cryptic lineages circulating in wastewaters ^34,35^ long before they spread in the population at large.

In support of the hypothesis that VOCs may emerge during prolonged infections, a number of studies have shown elevated evolutionary rates driven by selection during chronic infections of immunocompromised individuals ^6–8^. Among the persistently infected individuals we identified, we also found strong evidence for positive selection and parallel evolution, particularly in Spike and ORF1ab. In the most extreme case, we observed one persistent infection with 33 substitutions over a four-month period, 20 of which were nonsynonymous.

Potentially more remarkable, however, was our discovery of viral infections exhibiting latent evolutionary dynamics, with no consensus level genome changes for two months or longer. As far as we are aware, this is the first study to demonstrate that decelerated evolutionary rates may be a common outcome of persistent infection.

The factors causing these low rates of evolution are unknown, but one possible explanation is infection of long-lived cells ^36,37^ which then seed new outbreaks within the infected individual weeks or months later. Although the relapsing viral load dynamics we found in many of the persistently infected individuals supports this hypothesis, we did not find any consistent pattern between the amount of viral divergence during infection and the lower viral load activity in these individuals.

Intriguingly, individuals with persistent infections report fewer symptoms later in a persistent infection compared to at their first positive sample, or remain asymptomatic throughout infection, but have more than 50% higher odds of Long Covid compared to a control group. Although the link between viral persistence and Long Covid may not be causal, these results suggest persistent infections could be contributing to the pathophysiology of Long Covid ^10,38^, as also evidenced by the observation of circulating SARS-CoV-2 S1 spike protein in a subset of patients with Long Covid months after first infection ^39^. The association between persistent infection and Long Covid does not imply that every persistent infection can lead to Long Covid, only 9% of persistently infected individuals reported having Long Covid, nor does it mean that all cases of Long Covid are due to a persistent infection.

Indeed, many other possible mechanisms have been suggested to contribute to Long Covid including autoimmunity/inflammation, organ damage, EBV reactivation, and micro thrombosis (see ref ^10^ for a recent review).

Taken together, our observations highlight the continuing importance of community-based genomic surveillance both to monitor the emergence and spread of new variants, but also to gain a fundamental understanding of the natural history and evolution of novel pathogens and their clinical implications for patients.

## Methods

### ONS COVID-19 Infection Survey

The ONS-CIS is a UK household-based surveillance study in which participant households are approached at random from address lists across the country to provide a representative sample of the population ^40^. All individuals aged two years and older from each household who provide written informed consent provide swab samples (taken by the participant or parent/carer for those under 12 years), regardless of symptoms, and complete a questionnaire at assessments, which occur weekly for the first month in the survey and then monthly. From 26 April 2020 to 31 July 2022, assessments were conducted by study workers visiting each household; from 14 July 2022 onwards assessments were remote, with swabs taken using kits posted to participants and returned by post or courier, and questionnaires completed online or by telephone. Positive swab samples with Ct≤30 were sent for sequencing (see below). For this analysis, we included data from 2nd November 2020 to 15th August 2022, spanning a period from the earliest Alpha to latest Omicron BA.2 sequences within the ONS-CIS dataset.

This work contains statistical data from ONS which is Crown Copyright. The use of the ONS statistical data in this work does not imply the endorsement of the ONS in relation to the interpretation or analysis of the statistical data. This work uses research datasets which may not exactly reproduce National Statistics aggregates.

### Sequencing

From December 2020 onwards sequencing was attempted on all positive samples with Ct≤30; before this date, sequencing was attempted in real-time wherever possible, with some additional retrospective sequencing of stored samples. The vast majority of samples were sequenced on Illumina Novaseq, with a small number using Oxford Nanopore GridION or MINION. One of two protocols were used: either the ARTIC amplicon protocol ^41^ with consensus FASTA sequence files generated using the ARTIC nextflow processing pipeline ^42^, or veSeq, an RNASeq protocol based on a quantitative targeted enrichment strategy ^19,43^ with consensus sequences produced using *shiver* ^44^. During our study period, we identified 94,943 individuals with a single sequence and 5,774 individuals with two or more sequences. Here, we only included sequences with ≥50% genome coverage.

### Identifying rare SNPs

An important criterion for determining whether two sequences from the same individual are from the same infection is whether they share a rare single nucleotide polymorphism (SNP). These are defined as SNPs that are shared by fewer than 400 sequences corresponding to each major lineage within the full ONS-CIS dataset (**Supplementary Figure 1**). The thresholds were chosen to maximise the number of persistent infections identified whilst minimising the number of false positives (see below). The major lineages we considered were Alpha (B.1.1.7), Delta (B.1.617.2), Omicron BA.1 and Omicron BA.2, including their sublineages. Approximately 92-98% of all sequences from the four major lineages had a rare SNP relative to the major-lineage population-level consensus.

### Identifying reinfections with the same major lineage

Any pair of sequences from the same individual, of the same major lineage, and at least 26 days apart were considered as candidate reinfections. Of these, pairs that had at least one nucleotide difference at the consensus level, and did not share any rare SNPs, were classed as reinfections. Pairs that had no identical rare SNPs, nor any nucleotide differences at the consensus level, were classed as undetermined.

### Identifying individuals with persistent infection

We first identified individuals with two or more sequenced samples taken at least 26 days apart. We chose this cutoff because the majority of acutely infected individuals shed the virus for <20 days and no longer than 30 days in the respiratory tract ^23,45^. Given the extreme heterogeneity in the shedding profiles during some acute infections ^23^, we also considered a more conservative 56-day cutoff for some analyses.

For each identified individual we calculated the number of consensus nucleotide differences per site for all within-individual pairwise combinations of samples (**Supplementary Figure 6**). Only sites where a nucleotide difference could be called were included.

Candidate persistent infections were defined in one of two ways: (1) pairs of sequenced samples that belonged to the same major lineage, and (2) pairs of sequenced samples where one or both had no defined phylogenetic lineage, but where the genetic distance between them was lower than that required to differentiate two major lineages (see **Supplementary Figure 6**). We assumed pairs belonging to different major lineages were either coinfections or reinfections with two different virus lineages. Only candidate persistent infections were considered in further analysis.

Among the pool of candidate persistent infections, we defined persistent infections as those with sequences sharing one or more rare SNPs at two or more consecutive time points relative to the population-level consensus. A rare SNP is one that is observed in 400 or fewer samples within the entire ONS-CIS dataset.

### Determining the false positive rate for persistent infections

For each major lineage we generated a data set of 1,000 randomly paired sequences from different individuals in the ONS-CIS, each sampled at least 26 days apart. We determined the number of these pairs that would have been incorrectly identified as persistent infections as a function of the threshold for determining if a SNP is rare (**Supplementary Figure 1**). Although the total number of persistent infections identified grew as the threshold for determining if a SNP is rare increased, at very high thresholds the rate of false positives was also high. In our study, we chose a threshold of 400 sequences (corresponding to all sequences of the same major lineage within the full ONS-CIS dataset) for all of the major lineages, giving a false positive rate (identifying an infection as persistent when it was not) of 0-3%.

### Estimating the prevalence of persistent infections

Within the ONS-CIS we identified 54 infections that lasted 60 days or more. Comparing this to the number of individuals that had sequenced samples belonging to Alpha, Delta, BA.1 or BA.2, which to a good approximation will be the number of infections of these variants, we identified approximately 54/77,561 (0.07%) infections as persistent for >= 60 days. Since the ONS-CIS is a representative sample of individuals from the general population, we can estimate the percentage of all SARS-CoV-2 infections that became persistent for two months or longer. At one extreme, if all persistent infections have sequenceable virus for only four days per month (assuming viral dynamics similar to one acute infection each month), only 14% of persistent infections would be detected through monthly sampling. Correcting for this, we would estimate the percentage of persistent infections in the general population to be 0.5% (0.07% / 0.14). At the other extreme, if we assume all persistent-chronic infections (70% of persistent infections; see Main text) are detectable through monthly sampling, and the rest have detectable virus for 4 days per month, then we estimate that 74% (70% + 0.14*30%) of persistent infections were identified, giving an estimate of 0.09% (0.07% / 0.74) infections being persistent in the general population.

### Phylogenetic analysis

For each of the four major lineages, we chose 600 consensus sequences with at least 95% coverage from the ONS-CIS dataset using weighted random sampling, with each sample of major lineage *i* collected in week *j* given a weight *1/n*_*ij*_, where *n*_*ij*_ is the number of sequences of major lineage *i* collected during week *j* ^25^. These sequences were added as a background set to the collection of all consensus sequences for samples from persistent infections and reinfections. Mapping of each sequence to the Wuhan-Hu-1 reference sequence was already performed by *shiver* and thus a full alignment for each of the four lineages could be constructed using only this.

Maximum likelihood phylogenetic trees were constructed using IQ-TREE 1.6.12 ^46^ using the GTR+gamma substitution model and the ultrafast bootstrap ^47^. Each tree was rooted using the collection dates of the samples and the heuristic residual mean square algorithm in TempEst ^48^. Visualisation used ggtree ^49^.

### Comparing viral load activities and symptoms

To quantify the changes in viral load activities during persistent infections, we compared Ct values at the last time point a sequence was obtained to when the first sequence was collected. Similarly, for reinfections, we compared the changes in Ct value between the primary infection and reinfection. We used a paired t-test to calculate p-values in both cases as the distribution of differences in Ct values were normally distributed for both persistent infections (W=0.99, p=0.28) and reinfections (W=0.99, p=0.78) as determined by the Shapiro-Wilk test ^50^.

We also tracked 12 symptoms consistently solicited from all participants at every assessment. Symptoms were fever, weakness/tiredness, diarrhoea, shortness of breath, headache, nausea/vomiting, sore throat, muscle ache, abdominal pain, cough, loss of smell, and loss of taste. At each follow-up assessment, participants were asked whether these 12 symptoms had been present in the past seven days. Symptom discontinuation was defined as the first occurrence of two successive follow-up visits without reporting symptoms. To compare symptom counts during persistent infections and reinfections, we used the paired Wilcoxon test as the distribution of symptom differences is not normally distributed (see **Figure 3d**).

### Long Covid analysis

From February 2021, at every assessment, participants were asked “would you describe yourself as having Long Covid, that is, you are still experiencing symptoms more than 4 weeks after you first had COVID-19, that are not explained by something else?”. When estimating Long Covid prevalence in this analysis, we considered the first assessment at least 12 weeks and at least 26 weeks after infection. Our control group comprised all individuals with a positive PCR test and Ct≤30, excluding the persistently infected individuals identified in this study, over the same time span as persistent infections. We also ensured the follow-up from the start of infection to first Long Covid response was similar between persistent infections and controls (see **Table 2**).

In calculating the odds ratio of Long Covid in persistently infected individuals relative to the control group, we accounted for confounding variables such as age at the last birthday, sex, Ct value, calendar date, area deprivation quintile group, presence of self-reported long-term health conditions (binary), vaccination status (unvaccinated or single-vaccinated, fully-vaccinated or booster-vaccinated 14-89 days ago, fully-vaccinated or booster-vaccinated 90-179 days ago, fully-vaccinated or booster-vaccinated ≥180 days ago), and days from first positive test to Long Covid follow-up response. All variables except the last one were defined at the time of the first positive test. Continuous variables (age, Ct value, calendar date, days to follow-up response) were modelled as restricted cubic splines with a single internal knot at the median of the distribution and boundary knots at the 5^th^ and 95^th^ percentiles.

Vaccination status was derived from a combination of CIS and NIMS data for participants in England, and CIS data alone for participants in Wales, Scotland and Northern Ireland.

We were unable to do the Long Covid analysis for the reinfection group due to the low number of participants in this cohort who reported new-onset Long Covid ≥12 weeks or ≥26 weeks after infections.

## Data Availability

All genomic data have been made publicly available as part of the COVID-19 Genomics UK (COG-UK) Consortium 51. All other data, excluding personal clinical information on participants, are available in the main text, supplementary materials, or our GitHub repository (https://github.com/mg878/ONS-CIS_analysis).

## Data and materials availability

All genomic data have been made publicly available as part of the COVID-19 Genomics UK (COG-UK) Consortium ^51^. All other data, excluding personal clinical information on participants, are available in the main text, supplementary materials, or our GitHub repository (https://github.com/mg878/ONS-CIS_analysis).

## Supplementary figures

**Supplementary Figure 1:**
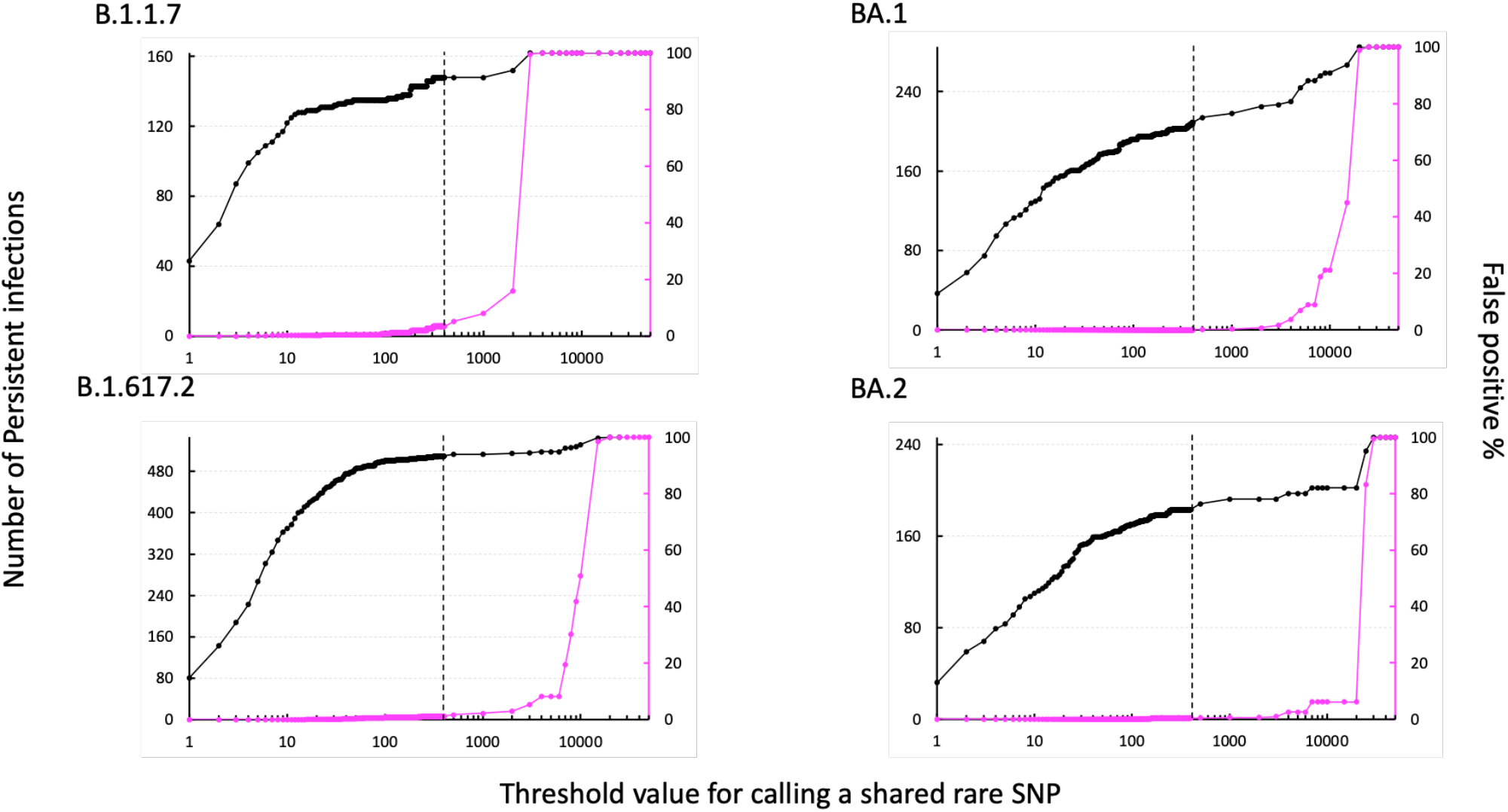
Number of persistent infections identified with a shared rare SNP as a function of the threshold for calling a rare SNP. Threshold value of 1 for rare SNPs implies the rare SNP is only found in one sequence of that lineage in the ONS-CIS dataset, excluding sequences from any persistently infected individuals. As the threshold value for calling a rare SNP increases, the number of persistent infections (of any duration) identified (black) increases until at some point the threshold value becomes so high that any individual with two or more sequences of the same major lineage would be identified as a persistent infection based on the rare SNP criterion at which point the false positivity rate (magenta) reaches 100%. At threshold value 400 (vertical dashed line) chosen in this study for identifying persistent infections, the percentage of false positives are 0% for BA.1 and BA.2 and 3% for Alpha and Delta.

**Supplementary Figure 2:**
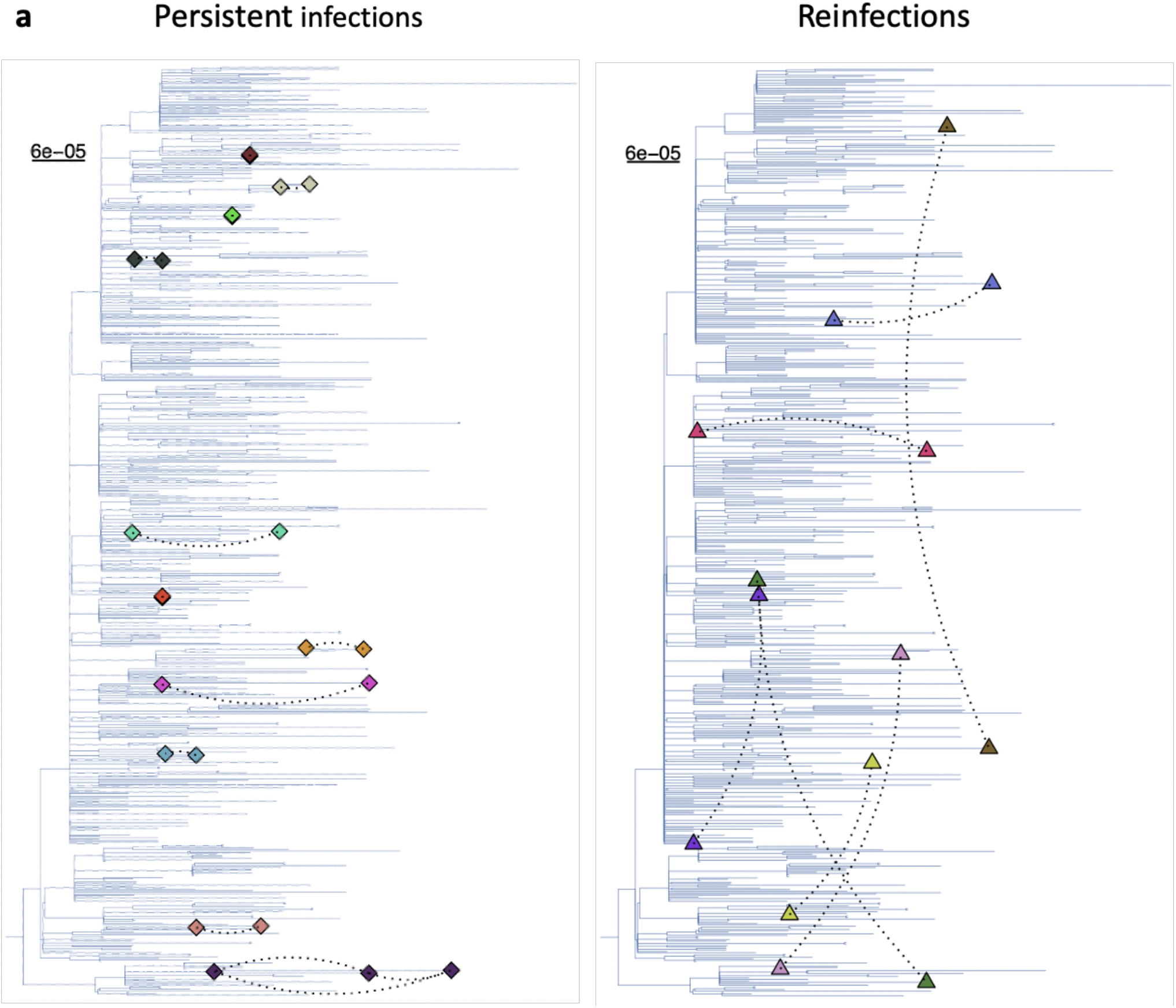
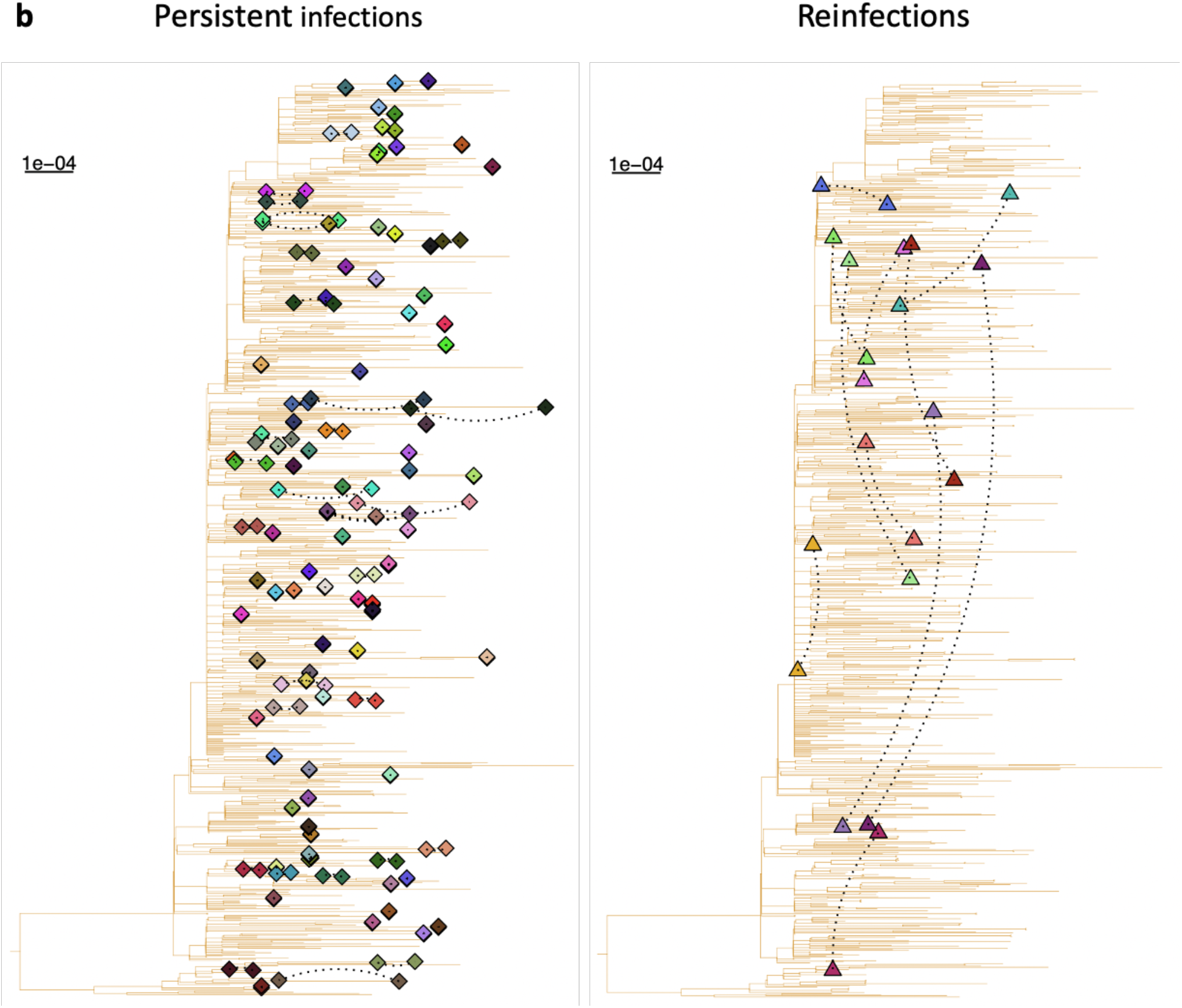
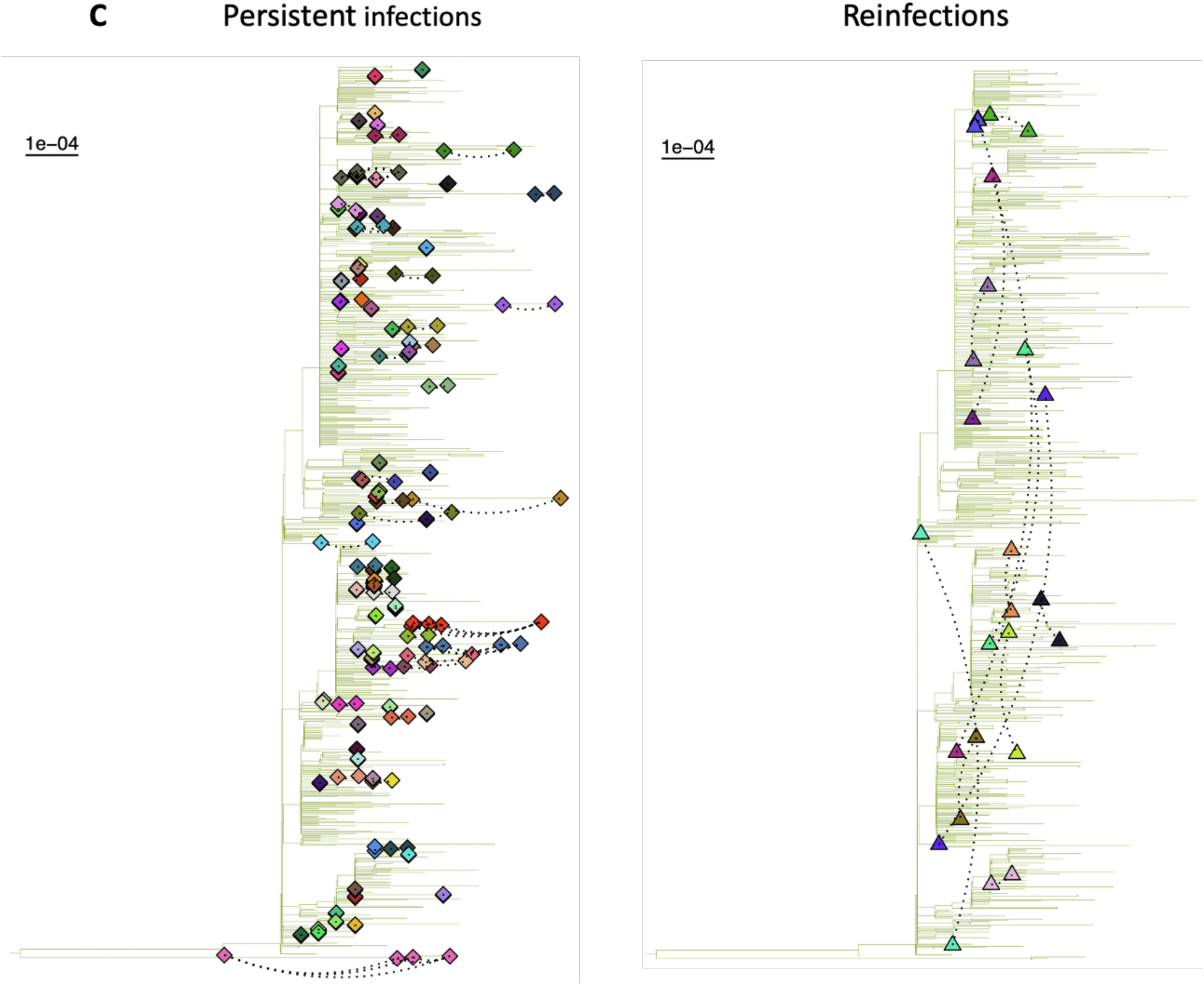
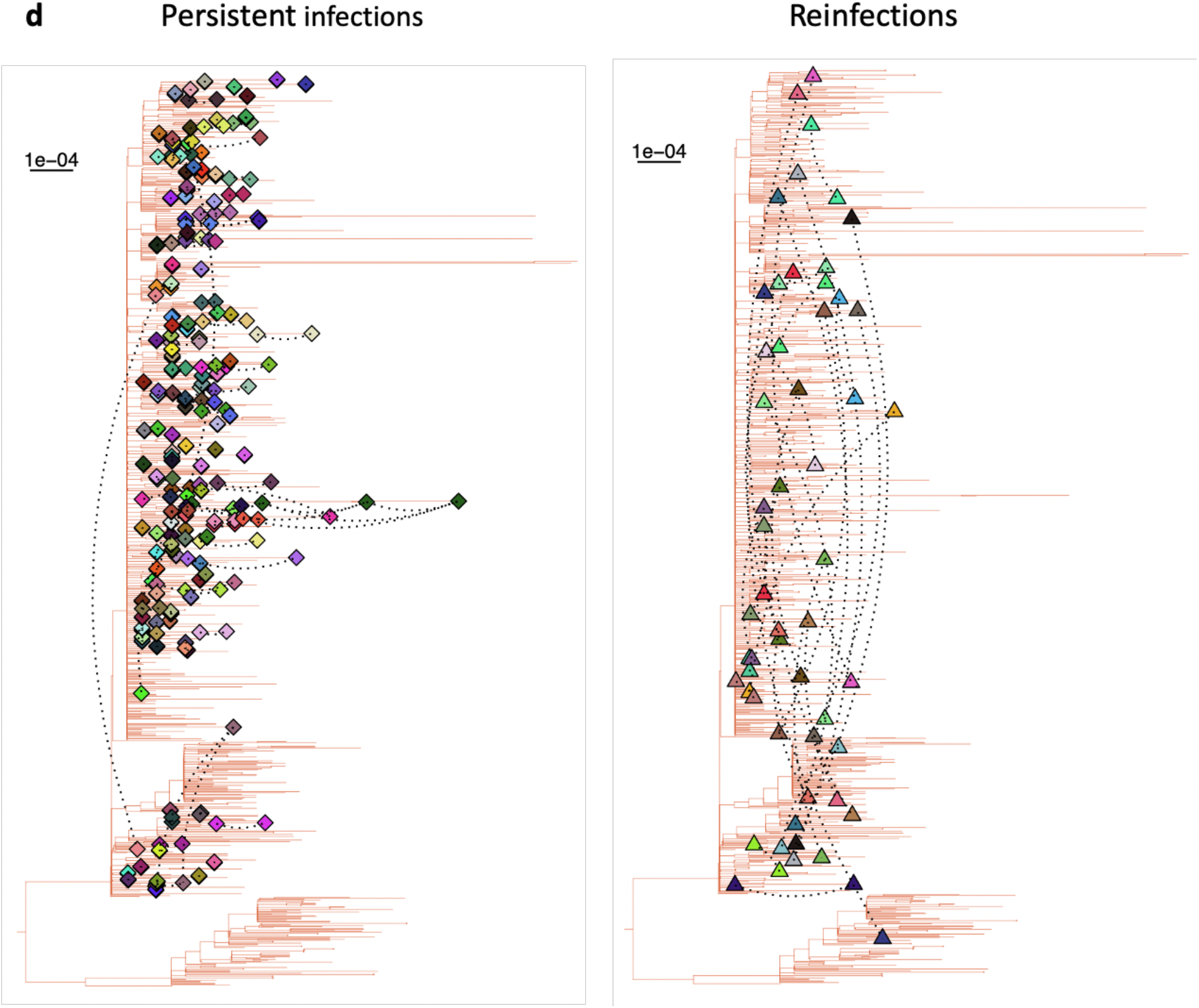
Phylogenetic relationship between samples from persistent infections and a representative background population per major lineage. Dashed lines connect every pair of sequences from the same individual. All sequences from the same individual are given the same colour. Pairs of sequences for (a) Alpha, (b) Delta, (c) Omicron BA.1, and (d) Omicron BA.2 that belong to persistent infections cluster closely together while reinfections do not. However, some of the sequences in 2 (out of 97) persistent infections with BA.1 and 5 (out of 167) persistent infections with BA.2 have poor bootstrap support (<80) and do not cluster together or cluster in a basal sister relationship. In all of these 7 cases, at least one of the sequences from each individual has a Ct value close to 30 with poor coverage. On the other hand, all sequences that belong to the same individual and have strong bootstrap support (>80) cluster together.

**Supplementary Figure 3:**
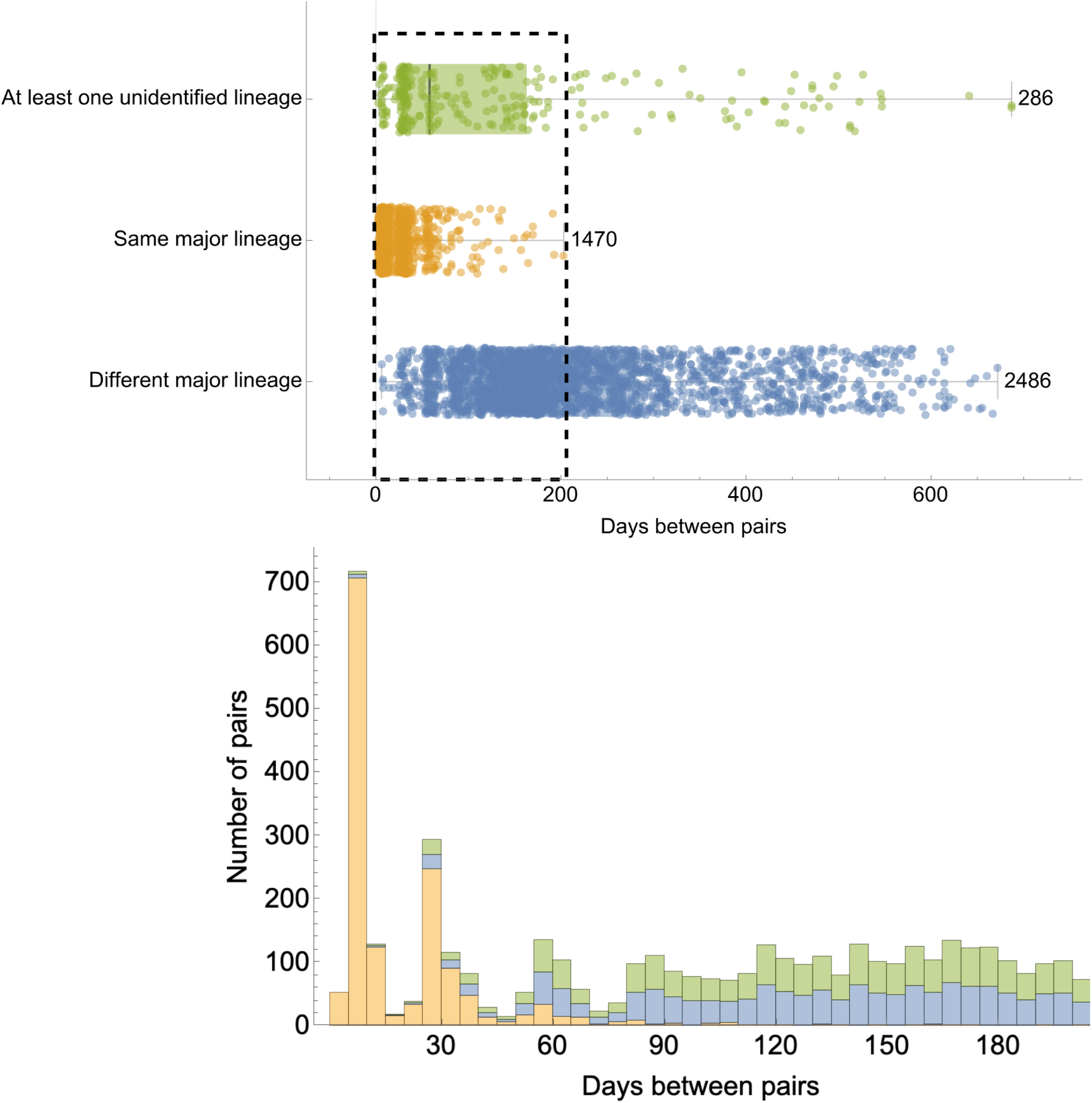
Days between all pairs of sequences from the same individual with two or more sequences. Pairs of sequences are classified as (i) pairs with at least one unidentified Pango lineage (green), (ii) pairs with identical major lineage (orange), and (iii) pairs from different major lineages. Bottom panel shows the counts of pairs in each of these three categories for the first 200-day time span (highlighted in a dashed rectangle in the top panel). Pairs include all possible combinations of sequences from the same individual. The number of pairs peaks at the 7-, 30-, and 60-day periods due to the sampling frequency of ONS-CIS (see **Methods**). Note that pairs with identical major lineage may not necessarily have identical Pango lineages (see **Methods**).

**Supplementary Figure 4:**
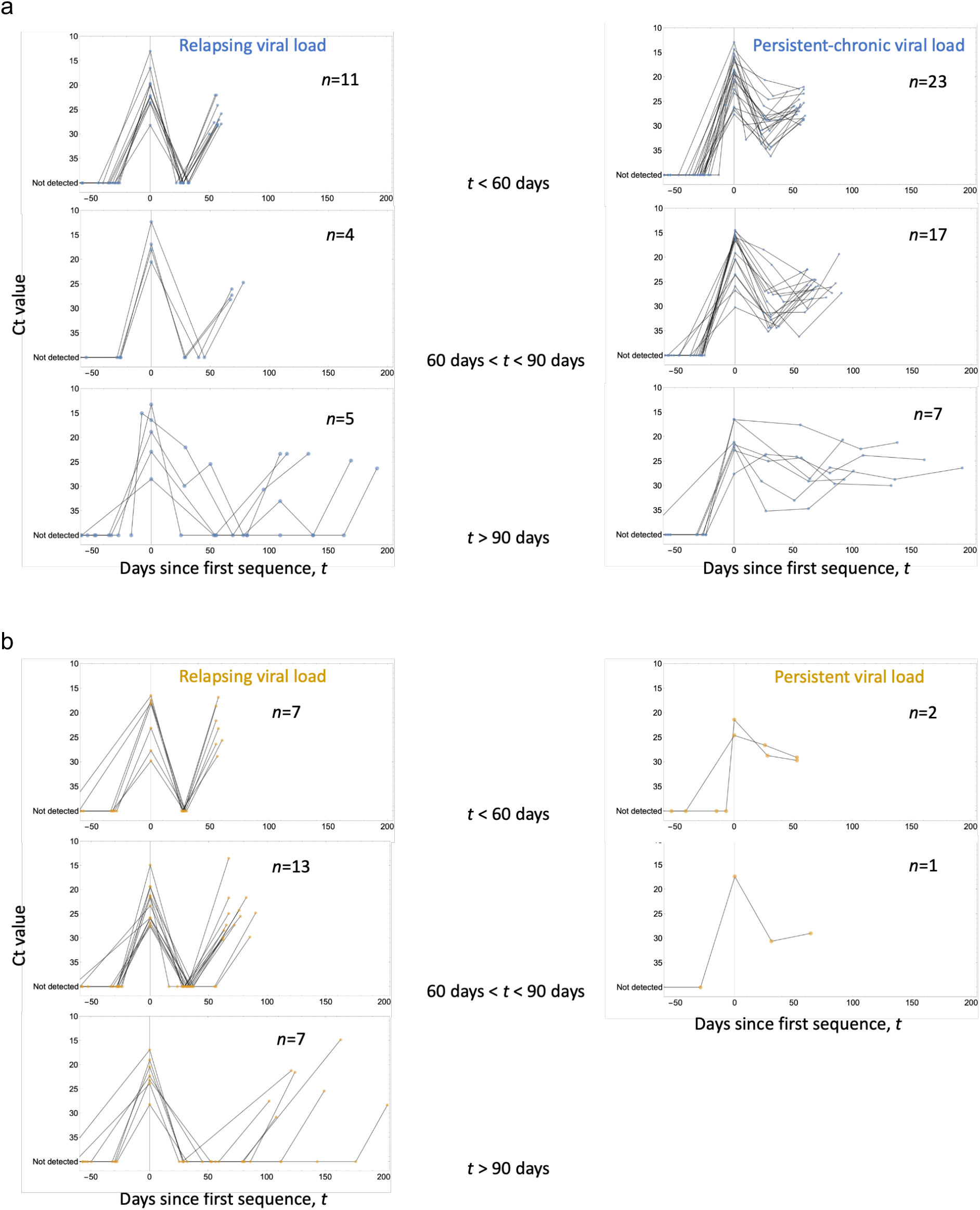
Viral load dynamics of individuals identified with persistent infections and reinfections stratified by duration and viral activity. Viral load activities of individuals, with 3 or more PCR tests taken during infection/until reinfection, identified as having **(a)** persistent infections and **(b)** reinfections with relapsing (left column) and persistent chronic (right column) trajectories. Three reinfections (two occurring in < 60 days and one between 60 to 90 days since first sequence) with persistent chronic viral load dynamics are excluded from the reinfection group as they are deemed potential persistent infections which do not have rare SNPs.

**Supplementary Figure 5:**
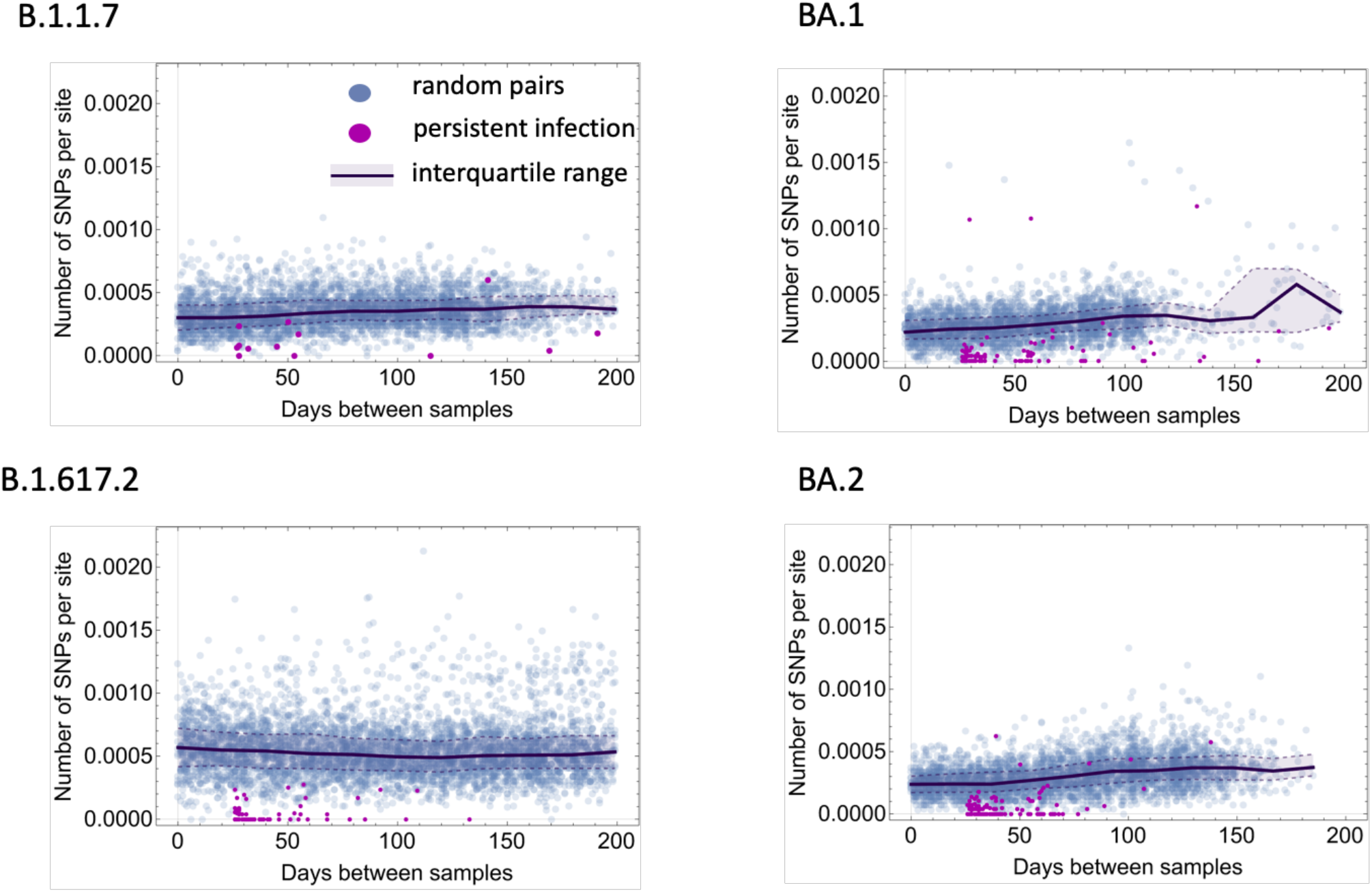
Number of single nucleotide polymorphisms detected in pairs of sequences from persistent infections vs. random pairs from a representative background population. Number of Single Nucleotide Variants (SNVs) per site between all the sequences collected from persistent infections (purple) and random pairs from individuals with only a single sequence within the ONS-CIS (blue) as a function of the number of days between each pair. For each major lineage, a pool of sequences from individuals with only one sequence within the ONS-CIS was sub-sampled and 500 random pairs generated for every 20 additional days between samples. For some major lineages where there were fewer than 500 pairs available beyond a certain time point, all possible random pairs within that 20-day period are used. Solid line and shaded area show the median and interquartile range, respectively, for random pairs over time. Note that the line and shaded area in each graph does not represent the rate of evolution but can be deemed as a measure of lineage diversity as a function of time difference between samples.

**Supplementary Figure 6:**
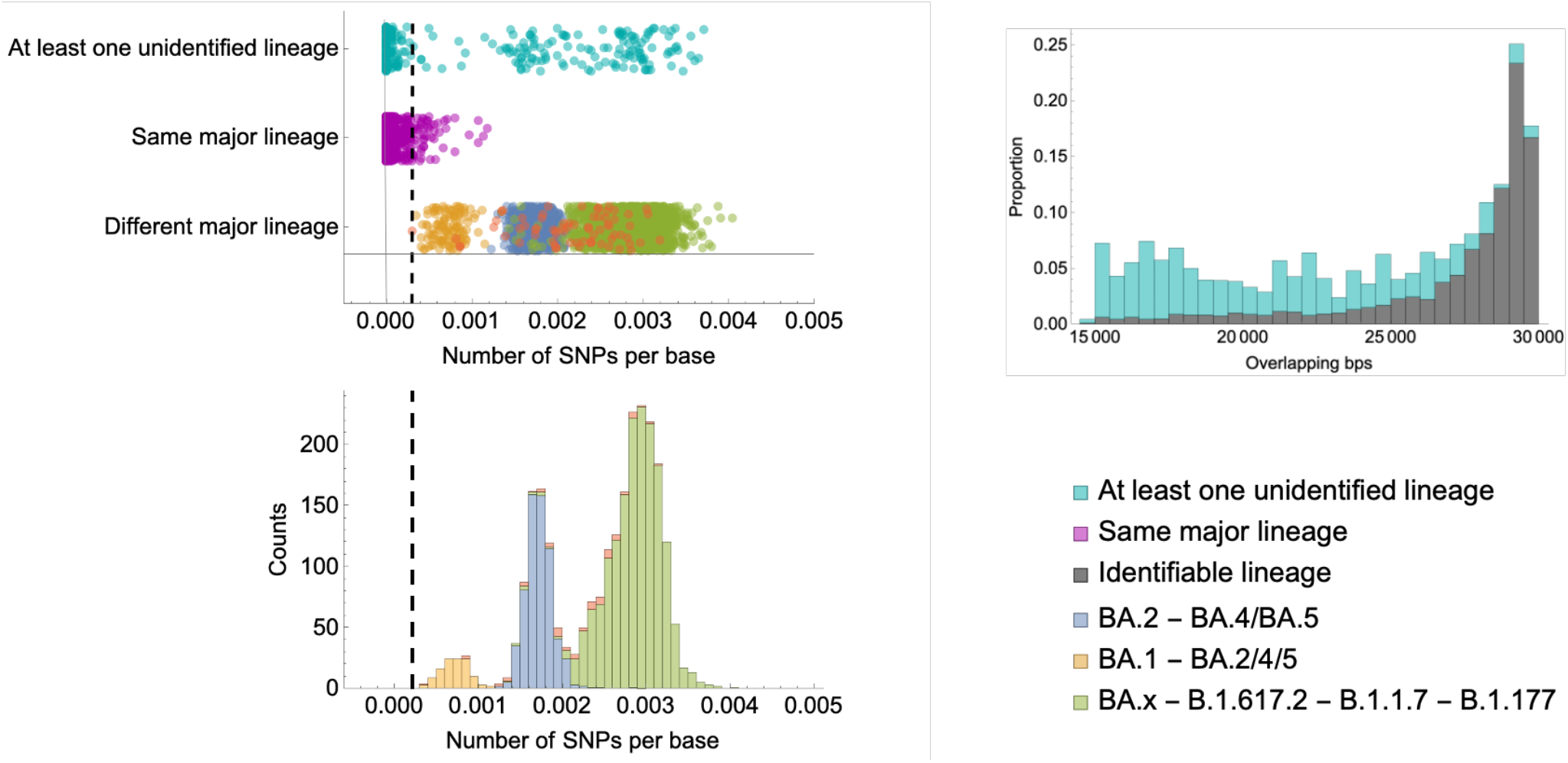
Pairwise differences between sequences from individuals with two or more sequences. (Left column) Number of single nucleotide polymorphisms (SNPs) between pairs of sequences from each individual with two or more sequences. Pairs include all possible combinations of sequences from the same individual. Vertical dashed line shows the lowest number of SNPs per base for pairs with different major lineages. Any pair with at least one unidentified lineage with a SNP per base smaller than the dashed line is selected as a candidate pair from a persistent infection. Pairs with different major lineages are coloured based on their number of SNPs per base into three groups: (i) pairs with one BA.1 and one BA.2 or BA.4 or BA.5 sequence (orange); (ii) pairs with one BA.2 and one BA.4 or BA.5 sequence (blue); and (iii) pairs with one Omicron (including all BA.x lineages) and one Delta (B.1.617.2), Alpha (B.1.1.7), or B.1.177 sequence (green). (Right column) Proportion of sequences with different number overlapping base pairs. Those with at least one unidentified lineage have a lower number of overlapping base pairs relative to pairs with identifiable lineage (i.e. pairs with identical or different major lineage) mainly due to having lower coverage.

## The COVID-19 Genomics UK (COG-UK) consortium

**June 2021 V.1**

Funding acquisition, Leadership and supervision, Metadata curation, Project administration, Samples and logistics, Sequencing and analysis, Software and analysis tools, and Visualisation:

Dr Samuel C Robson PhD ^13, 84^

Funding acquisition, Leadership and supervision, Metadata curation, Project administration, Samples and logistics, Sequencing and analysis, and Software and analysis tools:

Dr Thomas R Connor PhD ^11, 74^ and Prof Nicholas J Loman PhD ^43^

Leadership and supervision, Metadata curation, Project administration, Samples and logistics, Sequencing and analysis, Software and analysis tools, and Visualisation: Dr Tanya Golubchik PhD ^5^

Funding acquisition, Leadership and supervision, Metadata curation, Samples and logistics, Sequencing and analysis, and Visualisation:

Dr Rocio T Martinez Nunez PhD ^46^

Funding acquisition, Leadership and supervision, Project administration, Samples and logistics, Sequencing and analysis, and Software and analysis tools:

Dr David Bonsall PhD ^5^

Funding acquisition, Leadership and supervision, Project administration, Sequencing and analysis, Software and analysis tools, and Visualisation:

Prof Andrew Rambaut DPhil ^104^

Funding acquisition, Metadata curation, Project administration, Samples and logistics, Sequencing and analysis, and Software and analysis tools:

Dr Luke B Snell MSc, MBBS ^12^

Leadership and supervision, Metadata curation, Project administration, Samples and logistics, Software and analysis tools, and Visualisation:

Rich Livett MSc ^116^

Funding acquisition, Leadership and supervision, Metadata curation, Project administration, and Samples and logistics:

Dr Catherine Ludden PhD ^20, 70^

Funding acquisition, Leadership and supervision, Metadata curation, Samples and logistics, and Sequencing and analysis:

Dr Sally Corden PhD ^74^ and Dr Eleni Nastouli FRCPath ^96, 95, 30^

Funding acquisition, Leadership and supervision, Metadata curation, Sequencing and analysis, and Software and analysis tools:

Dr Gaia Nebbia PhD, FRCPath ^12^

Funding acquisition, Leadership and supervision, Project administration, Samples and logistics, and Sequencing and analysis:

Ian Johnston BSc ^116^

Leadership and supervision, Metadata curation, Project administration, Samples and logistics, and Sequencing and analysis:

Prof Katrina Lythgoe PhD ^5^, Dr M. Estee Torok FRCP ^19, 20^ and Prof Ian G Goodfellow PhD ^24^ Leadership and supervision, Metadata curation, Project administration, Samples and logistics, and Visualisation:

Dr Jacqui A Prieto PhD ^97, 82^ and Dr Kordo Saeed MD, FRCPath ^97, 83^

Leadership and supervision, Metadata curation, Project administration, Sequencing and analysis, and Software and analysis tools:

Dr David K Jackson PhD ^116^

Leadership and supervision, Metadata curation, Samples and logistics, Sequencing and analysis, and Visualisation:

Dr Catherine Houlihan PhD ^96, 94^

Leadership and supervision, Metadata curation, Sequencing and analysis, Software and analysis tools, and Visualisation:

Dr Dan Frampton PhD ^94, 95^

Metadata curation, Project administration, Samples and logistics, Sequencing and analysis, and Software and analysis tools:

Dr William L Hamilton PhD ^19^ and Dr Adam A Witney PhD ^41^

Funding acquisition, Samples and logistics, Sequencing and analysis, and Visualisation: Dr Giselda Bucca PhD ^101^

Funding acquisition, Leadership and supervision, Metadata curation, and Project administration:

Dr Cassie F Pope PhD^40, 41^

Funding acquisition, Leadership and supervision, Metadata curation, and Samples and logistics:

Dr Catherine Moore PhD ^74^

Funding acquisition, Leadership and supervision, Metadata curation, and Sequencing and analysis:

Prof Emma C Thomson PhD, FRCP ^53^

Funding acquisition, Leadership and supervision, Project administration, and Samples and logistics:

Dr Ewan M Harrison PhD ^116, 102^

Funding acquisition, Leadership and supervision, Sequencing and analysis, and

Visualisation:

Prof Colin P Smith PhD ^101^

Leadership and supervision, Metadata curation, Project administration, and Sequencing and analysis:

Fiona Rogan BSc ^77^

Leadership and supervision, Metadata curation, Project administration, and Samples and logistics:

Shaun M Beckwith MSc ^6^, Abigail Murray Degree ^6^, Dawn Singleton HNC ^6^, Dr Kirstine Eastick PhD, FRCPath ^37^, Dr Liz A Sheridan PhD ^98^, Paul Randell MSc, PgD ^99^, Dr Leigh M Jackson PhD ^105^, Dr Cristina V Ariani PhD ^116^ and Dr Sónia Gonçalves PhD ^116^

Leadership and supervision, Metadata curation, Samples and logistics, and Sequencing and analysis:

Dr Derek J Fairley PhD ^3, 77^, Prof Matthew W Loose PhD ^18^ and Joanne Watkins MSc ^74^

Leadership and supervision, Metadata curation, Samples and logistics, and Visualisation: Dr Samuel Moses MD ^25, 106^

Leadership and supervision, Metadata curation, Sequencing and analysis, and Software and analysis tools:

Dr Sam Nicholls PhD ^43^, Dr Matthew Bull PhD ^74^ and Dr Roberto Amato PhD ^116^

Leadership and supervision, Project administration, Samples and logistics, and Sequencing and analysis:

Prof Darren L Smith PhD ^36, 65, 66^

Leadership and supervision, Sequencing and analysis, Software and analysis tools, and Visualisation:

Prof David M Aanensen PhD ^14, 116^ and Dr Jeffrey C Barrett PhD ^116^

Metadata curation, Project administration, Samples and logistics, and Sequencing and analysis:

Dr Dinesh Aggarwal MRCP^20, 116, 70^, Dr James G Shepherd MBCHB, MRCP ^53^, Dr Martin D Curran PhD ^71^ and Dr Surendra Parmar PhD ^71^

Metadata curation, Project administration, Sequencing and analysis, and Software and analysis tools:

Dr Matthew D Parker PhD ^109^

Metadata curation, Samples and logistics, Sequencing and analysis, and Software and analysis tools:

Dr Catryn Williams PhD ^74^

Metadata curation, Samples and logistics, Sequencing and analysis, and Visualisation: Dr Sharon Glaysher PhD ^68^

Metadata curation, Sequencing and analysis, Software and analysis tools, and Visualisation: Dr Anthony P Underwood PhD ^14, 116^, Dr Matthew Bashton PhD ^36, 65^, Dr Nicole Pacchiarini PhD ^74^, Dr Katie F Loveson PhD ^84^ and Matthew Byott MSc ^95, 96^

Project administration, Sequencing and analysis, Software and analysis tools, and Visualisation:

Dr Alessandro M Carabelli PhD ^20^

Funding acquisition, Leadership and supervision, and Metadata curation: Dr Kate E Templeton PhD ^56, 104^

Funding acquisition, Leadership and supervision, and Project administration:

Dr Thushan I de Silva PhD ^109^, Dr Dennis Wang PhD ^109^, Dr Cordelia F Langford PhD ^116^ and John Sillitoe BEng ^116^

Funding acquisition, Leadership and supervision, and Samples and logistics: Prof Rory N Gunson PhD, FRCPath ^55^

Funding acquisition, Leadership and supervision, and Sequencing and analysis:

Dr Simon Cottrell PhD ^74^, Dr Justin O’Grady PhD ^75, 103^ and Prof Dominic Kwiatkowski PhD^116, 108^

Leadership and supervision, Metadata curation, and Project administration: Dr Patrick J Lillie PhD, FRCP ^37^

Leadership and supervision, Metadata curation, and Samples and logistics:

Dr Nicholas Cortes MBCHB ^33^, Dr Nathan Moore MBCHB ^33^, Dr Claire Thomas DPhil ^33^, Phillipa J Burns MSc, DipRCPath ^37^, Dr Tabitha W Mahungu FRCPath ^80^ and Steven Liggett BSc ^86^

Leadership and supervision, Metadata curation, and Sequencing and analysis: Angela H Beckett MSc ^13, 81^ and Prof Matthew TG Holden PhD ^73^

Leadership and supervision, Project administration, and Samples and logistics:

Dr Lisa J Levett PhD ^34^, Dr Husam Osman PhD ^70, 35^ and Dr Mohammed O Hassan-Ibrahim PhD, FRCPath ^99^

Leadership and supervision, Project administration, and Sequencing and analysis: Dr David A Simpson PhD ^77^

Leadership and supervision, Samples and logistics, and Sequencing and analysis:

Dr Meera Chand PhD ^72^, Prof Ravi K Gupta PhD ^102^, Prof Alistair C Darby PhD ^107^ and Prof Steve Paterson PhD ^107^

Leadership and supervision, Sequencing and analysis, and Software and analysis tools: Prof Oliver G Pybus DPhil ^23^, Dr Erik M Volz PhD ^39^, Prof Daniela de Angelis PhD ^52^, Prof David L Robertson PhD ^53^, Dr Andrew J Page PhD ^75^ and Dr Inigo Martincorena PhD ^116^

Leadership and supervision, Sequencing and analysis, and Visualisation: Dr Louise Aigrain PhD ^116^ and Dr Andrew R Bassett PhD ^116^

Metadata curation, Project administration, and Samples and logistics:

Dr Nick Wong DPhil, MRCP, FRCPath ^50^, Dr Yusri Taha MD, PhD ^89^, Michelle J Erkiert BA ^99^ and Dr Michael H Spencer Chapman MBBS ^116, 102^

Metadata curation, Project administration, and Sequencing and analysis: Dr Rebecca Dewar PhD ^56^ and Martin P McHugh MSc ^56, 111^

Metadata curation, Project administration, and Software and analysis tools: Siddharth Mookerjee MPH ^38, 57^

Metadata curation, Project administration, and Visualisation:

Stephen Aplin ^97^, Matthew Harvey ^97^, Thea Sass ^97^, Dr Helen Umpleby FRCP ^97^ and Helen Wheeler ^97^

Metadata curation, Samples and logistics, and Sequencing and analysis:

Dr James P McKenna PhD ^3^, Dr Ben Warne MRCP ^9^, Joshua F Taylor MSc ^22^, Yasmin Chaudhry BSc ^24^, Rhys Izuagbe ^24^, Dr Aminu S Jahun PhD ^24^, Dr Gregory R Young PhD ^36, 65^, Dr Claire McMurray PhD ^43^, Dr Clare M McCann PhD ^65, 66^, Dr Andrew Nelson PhD ^65, 66^ and Scott Elliott ^68^

Metadata curation, Samples and logistics, and Visualisation:

Hannah Lowe MSc ^25^

Metadata curation, Sequencing and analysis, and Software and analysis tools:

Dr Anna Price PhD ^11^, Matthew R Crown BSc ^65^, Dr Sara Rey PhD ^74^, Dr Sunando Roy PhD

^96^ and Dr Ben Temperton PhD ^105^

Metadata curation, Sequencing and analysis, and Visualisation:

Dr Sharif Shaaban PhD ^73^ and Dr Andrew R Hesketh PhD ^101^

Project administration, Samples and logistics, and Sequencing and analysis:

Dr Kenneth G Laing PhD^41^, Dr Irene M Monahan PhD ^41^ and Dr Judith Heaney PhD ^95, 96, 34^

Project administration, Samples and logistics, and Visualisation:

Dr Emanuela Pelosi FRCPath ^97^, Siona Silviera MSc ^97^ and Dr Eleri Wilson-Davies MD, FRCPath ^97^

Samples and logistics, Software and analysis tools, and Visualisation: Dr Helen Fryer PhD ^5^

Sequencing and analysis, Software and analysis tools, and Visualization:

Dr Helen Adams PhD ^4^, Dr Louis du Plessis PhD ^23^, Dr Rob Johnson PhD ^39^, Dr William T Harvey PhD ^53, 42^, Dr Joseph Hughes PhD ^53^, Dr Richard J Orton PhD ^53^, Dr Lewis G Spurgin PhD ^59^, Dr Yann Bourgeois PhD ^81^, Dr Chris Ruis PhD ^102^, Áine O’Toole MSc ^104^, Marina Gourtovaia MSc ^116^ and Dr Theo Sanderson PhD ^116^

Funding acquisition, and Leadership and supervision:

Dr Christophe Fraser PhD ^5^, Dr Jonathan Edgeworth PhD, FRCPath ^12^, Prof Judith Breuer MD ^96, 29^, Dr Stephen L Michell PhD ^105^ and Prof John A Todd PhD ^115^

Funding acquisition, and Project administration:

Michaela John BSc ^10^ and Dr David Buck PhD ^115^

Leadership and supervision, and Metadata curation:

Dr Kavitha Gajee MBBS, FRCPath ^37^ and Dr Gemma L Kay PhD ^75^

Leadership and supervision, and Project administration:

Prof Sharon J Peacock PhD ^20, 70^ and David Heyburn ^74^

Leadership and supervision, and Samples and logistics:

Katie Kitchman BSc ^37^, Prof Alan McNally PhD ^43, 93^, David T Pritchard MSc, CSci ^50^, Dr Samir Dervisevic FRCPath ^58^, Dr Peter Muir PhD ^70^, Dr Esther Robinson PhD ^70, 35^, Dr Barry B Vipond PhD ^70^, Newara A Ramadan MSc, CSci, FIBMS ^78^, Dr Christopher Jeanes MBBS ^90^, Danni Weldon BSc ^116^, Jana Catalan MSc ^118^ and Neil Jones MSc ^118^Leadership and supervision, and Sequencing and analysis: Dr Ana da Silva Filipe PhD ^53^, Dr Chris Williams MBBS ^74^, Marc Fuchs BSc ^77^, Dr Julia Miskelly PhD ^77^, Dr Aaron R Jeffries PhD ^105^, Karen Oliver BSc ^116^ and Dr Naomi R Park PhD ^116^Metadata curation, and Samples and logistics: Amy Ash BSc ^1^, Cherian Koshy MSc, CSci, FIBMS ^1^, Magdalena Barrow ^7^, Dr Sarah L Buchan PhD ^7^, Dr Anna Mantzouratou PhD ^7^, Dr Gemma Clark PhD ^15^, Dr Christopher W Holmes PhD ^16^, Sharon Campbell MSc ^17^, Thomas Davis MSc ^21^, Ngee Keong Tan MSc ^22^, Dr Julianne R Brown PhD ^29^, Dr Kathryn A Harris PhD ^29, 2^, Stephen P Kidd MSc ^33^, Dr Paul R Grant PhD ^34^, Dr Li Xu-McCrae PhD ^35^, Dr Alison Cox PhD ^38, 63^, Pinglawathee Madona ^38, 63^, Dr Marcus Pond PhD ^38, 63^, Dr Paul A Randell MBBCh ^38, 63^, Karen T Withell FIBMS ^48^, Cheryl Williams MSc ^51^, Dr Clive Graham MD ^60^, Rebecca Denton-Smith BSc ^62^, Emma Swindells BSc ^62^, Robyn Turnbull BSc ^62^, Dr Tim J Sloan PhD ^67^, Dr Andrew Bosworth PhD ^70, 35^, Stephanie Hutchings ^70^, Hannah M Pymont MSc ^70^, Dr Anna Casey PhD ^76^, Dr Liz Ratcliffe PhD ^76^, Dr Christopher R Jones PhD ^79, 105^, Dr Bridget A Knight PhD ^79, 105^, Dr Tanzina Haque PhD, FRCPath ^80^, Dr Jennifer Hart MRCP ^80^, Dr Dianne Irish-Tavares FRCPath ^80^, Eric Witele MSc ^80^, Craig Mower BA ^86^, Louisa K Watson DipHE ^86^, Jennifer Collins BSc ^89^, Gary Eltringham BSc ^89^, Dorian Crudgington ^98^, Ben Macklin ^98^, Prof Miren Iturriza-Gomara PhD ^107^, Dr Anita O Lucaci PhD ^107^ and Dr Patrick C McClure PhD ^113^

Metadata curation, and Sequencing and analysis:

Matthew Carlile BSc ^18^, Dr Nadine Holmes PhD ^18^, Dr Christopher Moore PhD ^18^, Dr Nathaniel Storey PhD ^29^, Dr Stefan Rooke PhD ^73^, Dr Gonzalo Yebra PhD ^73^, Dr Noel Craine DPhil ^74^, Malorie Perry MSc ^74^, Dr Nabil-Fareed Alikhan PhD ^75^, Dr Stephen Bridgett PhD ^77^, Kate F Cook MScR ^84^, Christopher Fearn MSc ^84^, Dr Salman Goudarzi PhD ^84^, Prof Ronan A Lyons MD ^88^, Dr Thomas Williams MD ^104^, Dr Sam T Haldenby PhD ^107^, Jillian Durham BSc ^116^ and Dr Steven Leonard PhD ^116^

Metadata curation, and Software and analysis tools:

Robert M Davies MA (Cantab) ^116^

Project administration, and Samples and logistics:

Dr Rahul Batra MD ^12^, Beth Blane BSc ^20^, Dr Moira J Spyer PhD ^30, 95, 96^, Perminder Smith MSc ^32, 112^, Mehmet Yavus ^85, 109^, Dr Rachel J Williams PhD ^96^, Dr Adhyana IK Mahanama MD ^97^, Dr Buddhini Samaraweera MD ^97^, Sophia T Girgis MSc ^102^, Samantha E Hansford CSci ^109^, Dr Angie Green PhD ^115^, Dr Charlotte Beaver PhD ^116^, Katherine L Bellis ^116, 102^, Matthew J Dorman ^116^, Sally Kay ^116^, Liam Prestwood ^116^ and Dr Shavanthi Rajatileka PhD ^116^

Project administration, and Sequencing and analysis:

Dr Joshua Quick PhD ^43^

Project administration, and Software and analysis tools:

Radoslaw Poplawski BSc ^43^

Samples and logistics, and Sequencing and analysis:

Dr Nicola Reynolds PhD ^8^, Andrew Mack MPhil ^11^, Dr Arthur Morriss PhD ^11^, Thomas Whalley BSc ^11^, Bindi Patel BSc ^12^, Dr Iliana Georgana PhD ^24^, Dr Myra Hosmillo PhD ^24^, Malte L Pinckert MPhil ^24^, Dr Joanne Stockton PhD ^43^, Dr John H Henderson PhD ^65^, Amy Hollis HND ^65^, Dr William Stanley PhD ^65^, Dr Wen C Yew PhD ^65^, Dr Richard Myers PhD ^72^, Dr Alicia Thornton PhD ^72^, Alexander Adams BSc ^74^, Tara Annett BSc ^74^, Dr Hibo Asad PhD ^74^, Alec Birchley MSc ^74^, Jason Coombes BSc ^74^, Johnathan M Evans MSc ^74^, Laia Fina ^74^, Bree Gatica-Wilcox MPhil ^74^, Lauren Gilbert ^74^, Lee Graham BSc ^74^, Jessica Hey BSc ^74^, Ember Hilvers MPH ^74^, Sophie Jones MSc ^74^, Hannah Jones ^74^, Sara Kumziene-Summerhayes MSc ^74^, Dr Caoimhe McKerr PhD ^74^, Jessica Powell BSc ^74^, Georgia Pugh ^74^, Sarah Taylor ^74^, Alexander J Trotter MRes ^75^, Charlotte A Williams BSc ^96^, Leanne M Kermack MSc ^102^, Benjamin H Foulkes MSc ^109^, Marta Gallis MSc ^109^, Hailey R Hornsby MSc ^109^, Stavroula F Louka MSc ^109^, Dr Manoj Pohare PhD ^109^, Paige Wolverson MSc ^109^, Peijun Zhang MSc ^109^, George MacIntyre-Cockett BSc ^115^, Amy Trebes MSc ^115^, Dr Robin J Moll PhD ^116^, Lynne Ferguson MSc ^117^, Dr Emily J Goldstein PhD ^117^, Dr Alasdair Maclean PhD ^117^ and Dr Rachael Tomb PhD ^117^

Samples and logistics, and Software and analysis tools:

Dr Igor Starinskij MSc, MRCP ^53^

Sequencing and analysis, and Software and analysis tools:

Laura Thomson BSc ^5^, Joel Southgate MSc ^11, 74^, Dr Moritz UG Kraemer DPhil ^23^, Dr Jayna Raghwani PhD ^23^, Dr Alex E Zarebski PhD ^23^, Olivia Boyd MSc ^39^, Lily Geidelberg MSc ^39^, Dr Chris J Illingworth PhD ^52^, Dr Chris Jackson PhD ^52^, Dr David Pascall PhD ^52^, Dr Sreenu Vattipally PhD ^53^, Timothy M Freeman MPhil ^109^, Dr Sharon N Hsu PhD ^109^, Dr Benjamin B Lindsey MRCP ^109^, Dr Keith James PhD ^116^, Kevin Lewis ^116^, Gerry Tonkin-Hill ^116^ and Dr Jaime M Tovar-Corona PhD ^116^

Sequencing and analysis, and Visualisation:

MacGregor Cox MSci ^20^

Software and analysis tools, and Visualisation:

Dr Khalil Abudahab PhD ^14, 116^, Mirko Menegazzo ^14^, Ben EW Taylor MEng ^14, 116^, Dr Corin A Yeats PhD ^14^, Afrida Mukaddas BTech ^53^, Derek W Wright MSc ^53^, Dr Leonardo de Oliveira Martins PhD ^75^, Dr Rachel Colquhoun DPhil ^104^, Verity Hill ^104^, Dr Ben Jackson PhD ^104^, Dr JT McCrone PhD ^104^, Dr Nathan Medd PhD ^104^, Dr Emily Scher PhD ^104^ and Jon-Paul Keatley ^116^

Leadership and supervision:

Dr Tanya Curran PhD ^3^, Dr Sian Morgan FRCPath ^10^, Prof Patrick Maxwell PhD ^20^, Prof Ken Smith PhD ^20^, Dr Sahar Eldirdiri MBBS, MSc, FRCPath ^21^, Anita Kenyon MSc ^21^, Prof Alison H Holmes MD ^38, 57^, Dr James R Price PhD ^38, 57^, Dr Tim Wyatt PhD ^69^, Dr Alison E Mather PhD ^75^, Dr Timofey Skvortsov PhD ^77^ and Prof John A Hartley PhD ^96^

Metadata curation:

Prof Martyn Guest PhD ^11^, Dr Christine Kitchen PhD ^11^, Dr Ian Merrick PhD ^11^, Robert Munn BSc ^11^, Dr Beatrice Bertolusso Degree ^33^, Dr Jessica Lynch MBCHB ^33^, Dr Gabrielle Vernet MBBS ^33^, Stuart Kirk MSc ^34^, Dr Elizabeth Wastnedge MD ^56^, Dr Rachael Stanley PhD ^58^, Giles Idle ^64^, Dr Declan T Bradley PhD ^69, 77^, Dr Jennifer Poyner MD ^79^ and Matilde Mori BSc ^110^

Project administration:

Owen Jones BSc ^11^, Victoria Wright BSc ^18^, Ellena Brooks MA ^20^, Carol M Churcher BSc ^20^, Mireille Fragakis HND ^20^, Dr Katerina Galai PhD ^20, 70^, Dr Andrew Jermy PhD ^20^, Sarah Judges BA ^20^, Georgina M McManus BSc ^20^, Kim S Smith ^20^, Dr Elaine Westwick PhD ^20^, Dr Stephen W Attwood PhD ^23^, Dr Frances Bolt PhD ^38, 57^, Dr Alisha Davies PhD ^74^, Elen De Lacy MPH ^74^, Fatima Downing ^74^, Sue Edwards ^74^, Lizzie Meadows MA ^75^, Sarah Jeremiah MSc ^97^, Dr Nikki Smith PhD ^109^ and Luke Foulser ^116^

Samples and logistics:

Dr Themoula Charalampous PhD ^12, 46^, Amita Patel BSc ^12^, Dr Louise Berry PhD ^15^, Dr Tim Boswell PhD ^15^, Dr Vicki M Fleming PhD ^15^, Dr Hannah C Howson-Wells PhD ^15^, Dr Amelia Joseph PhD ^15^, Manjinder Khakh ^15^, Dr Michelle M Lister PhD ^15^, Paul W Bird MSc, MRes ^16^, Karlie Fallon ^16^, Thomas Helmer ^16^, Dr Claire L McMurray PhD ^16^, Mina Odedra BSc ^16^, Jessica Shaw BSc ^16^, Dr Julian W Tang PhD ^16^, Nicholas J Willford MSc ^16^, Victoria Blakey BSc ^17^, Dr Veena Raviprakash MD ^17^, Nicola Sheriff BSc ^17^, Lesley-Anne Williams BSc ^17^, Theresa Feltwell MSc ^20^, Dr Luke Bedford PhD ^26^, Dr James S Cargill PhD ^27^, Warwick Hughes MSc ^27^, Dr Jonathan Moore MD ^28^, Susanne Stonehouse BSc ^28^, Laura Atkinson MSc ^29^, Jack CD Lee MSc ^29^, Dr Divya Shah PhD ^29^, Adela Alcolea-Medina Clinical scientist ^32, 112^, Natasha Ohemeng-Kumi MSc ^32, 112^, John Ramble MSc ^32, 112^, Jasveen Sehmi MSc ^32, 112^, Dr Rebecca Williams BMBS ^33^, Wendy Chatterton MSc ^34^, Monika Pusok MSc ^34^, William Everson MSc ^37^, Anibolina Castigador IBMS HCPC ^44^, Emily Macnaughton FRCPath ^44^, Dr Kate El Bouzidi MRCP ^45^, Dr Temi Lampejo FRCPath ^45^, Dr Malur Sudhanva FRCPath ^45^, Cassie Breen BSc ^47^, Dr Graciela Sluga MD, MSc ^48^, Dr Shazaad SY Ahmad MSc ^49, 70^, Dr Ryan P George PhD ^49^, Dr Nicholas W Machin MSc ^49, 70^, Debbie Binns BSc ^50^, Victoria James BSc ^50^, Dr Rachel Blacow MBCHB ^55^, Dr Lindsay Coupland PhD ^58^, Dr Louise Smith PhD ^59^, Dr Edward Barton MD ^60^, Debra Padgett BSc ^60^, Garren Scott BSc ^60^, Dr Aidan Cross MBCHB ^61^, Dr Mariyam Mirfenderesky FRCPath ^61^, Jane Greenaway MSc ^62^, Kevin Cole ^64^, Phillip Clarke ^67^, Nichola Duckworth ^67^, Sarah Walsh ^67^, Kelly Bicknell ^68^, Robert Impey MSc ^68^, Dr Sarah Wyllie PhD ^68^, Richard Hopes ^70^, Dr Chloe Bishop PhD ^72^, Dr Vicki Chalker PhD ^72^, Dr Ian Harrison PhD ^72^, Laura Gifford MSc ^74^, Dr Zoltan Molnar PhD ^77^, Dr Cressida Auckland FRCPath ^79^, Dr Cariad Evans PhD ^85, 109^, Dr Kate Johnson PhD ^85, 109^, Dr David G Partridge FRCP, FRCPath ^85, 109^, Dr Mohammad Raza PhD ^85, 109^, Paul Baker MD ^86^, Prof Stephen Bonner PhD ^86^, Sarah Essex ^86^, Leanne J Murray ^86^, Andrew I Lawton MSc ^87^, Dr Shirelle Burton-Fanning MD ^89^, Dr Brendan AI Payne MD ^89^, Dr Sheila Waugh MD ^89^, Andrea N Gomes MSc ^91^, Maimuna Kimuli MSc ^91^, Darren R Murray MSc ^91^, Paula Ashfield MSc ^92^, Dr Donald Dobie MBCHB ^92^, Dr Fiona Ashford PhD ^93^, Dr Angus Best PhD ^93^, Dr Liam Crawford PhD ^93^, Dr Nicola Cumley PhD ^93^, Dr Megan Mayhew PhD ^93^, Dr Oliver Megram PhD ^93^, Dr Jeremy Mirza PhD ^93^, Dr Emma Moles-Garcia PhD ^93^, Dr Benita Percival PhD ^93^, Megan Driscoll BSc ^96^, Leah Ensell BSc ^96^, Dr Helen L Lowe PhD ^96^, Laurentiu Maftei BSc ^96^, Matteo Mondani MSc ^96^, Nicola J Chaloner BSc ^99^, Benjamin J Cogger BSc ^99^, Lisa J Easton MSc ^99^, Hannah Huckson BSc ^99^, Jonathan Lewis MSc, PgD, FIBMS ^99^, Sarah Lowdon BSc ^99^, Cassandra S Malone MSc ^99^, Florence Munemo BSc ^99^, Manasa Mutingwende MSc ^99^, Roberto Nicodemi BSc ^99^, Olga Podplomyk FD ^99^, Thomas Somassa BSc ^99^, Dr Andrew Beggs PhD ^100^, Dr Alex Richter PhD ^100^, Claire Cormie ^102^, Joana Dias MSc ^102^, Sally Forrest BSc ^102^, Dr Ellen E Higginson PhD ^102^, Mailis Maes MPhil ^102^, Jamie Young BSc ^102^, Dr Rose K Davidson PhD ^103^, Kathryn A Jackson MSc ^107^, Dr Lance Turtle PhD, MRCP ^107^, Dr Alexander J Keeley MRCP ^109^, Prof Jonathan Ball PhD ^113^, Timothy Byaruhanga MSc ^113^, Dr Joseph G Chappell PhD ^113^, Jayasree Dey MSc ^113^, Jack D Hill MSc ^113^, Emily J Park MSc ^113^, Arezou Fanaie MSc ^114^, Rachel A Hilson MSc ^114^, Geraldine Yaze MSc ^114^ and Stephanie Lo ^116^

Sequencing and analysis:

Safiah Afifi BSc ^10^, Robert Beer BSc ^10^, Joshua Maksimovic FD ^10^, Kathryn McCluggage Masters ^10^, Karla Spellman FD ^10^, Catherine Bresner BSc ^11^, William Fuller BSc ^11^, Dr Angela Marchbank BSc ^11^, Trudy Workman HNC ^11^, Dr Ekaterina Shelest PhD ^13, 81^, Dr Johnny Debebe PhD ^18^, Dr Fei Sang PhD ^18^, Dr Marina Escalera Zamudio PhD ^23^, Dr Sarah Francois PhD ^23^, Bernardo Gutierrez MSc ^23^, Dr Tetyana I Vasylyeva DPhil ^23^, Dr Flavia Flaviani PhD ^31^, Dr Manon Ragonnet-Cronin PhD ^39^, Dr Katherine L Smollett PhD ^42^, Alice Broos BSc ^53^, Daniel Mair BSc ^53^, Jenna Nichols BSc ^53^, Dr Kyriaki Nomikou PhD ^53^, Dr Lily Tong PhD ^53^, Ioulia Tsatsani MSc ^53^, Prof Sarah O’Brien PhD ^54^, Prof Steven Rushton PhD ^54^, Dr Roy Sanderson PhD ^54^, Dr Jon Perkins MBCHB ^55^, Seb Cotton MSc ^56^, Abbie Gallagher BSc ^56^, Dr Elias Allara MD, PhD ^70, 102^, Clare Pearson MSc ^70, 102^, Dr David Bibby PhD ^72^, Dr Gavin Dabrera PhD ^72^, Dr Nicholas Ellaby PhD ^72^, Dr Eileen Gallagher PhD ^72^, Dr Jonathan Hubb PhD ^72^, Dr Angie Lackenby PhD ^72^, Dr David Lee PhD ^72^, Nikos Manesis ^72^, Dr Tamyo Mbisa PhD ^72^, Dr Steven Platt PhD ^72^, Katherine A Twohig ^72^, Dr Mari Morgan PhD ^74^, Alp Aydin MSci ^75^, David J Baker BEng ^75^, Dr Ebenezer Foster-Nyarko PhD ^75^, Dr Sophie J Prosolek PhD ^75^, Steven Rudder ^75^, Chris Baxter BSc ^77^, Sílvia F Carvalho MSc ^77^, Dr Deborah Lavin PhD ^77^, Dr Arun Mariappan PhD ^77^, Dr Clara Radulescu PhD ^77^, Dr Aditi Singh PhD ^77^, Miao Tang MD ^77^, Helen Morcrette BSc ^79^, Nadua Bayzid BSc ^96^, Marius Cotic MSc ^96^, Dr Carlos E Balcazar PhD ^104^, Dr Michael D Gallagher PhD ^104^, Dr Daniel Maloney PhD ^104^, Thomas D Stanton BSc ^104^, Dr Kathleen A Williamson PhD ^104^, Dr Robin Manley PhD ^105^, Michelle L Michelsen BSc ^105^, Dr Christine M Sambles PhD ^105^, Dr David J Studholme PhD ^105^, Joanna Warwick-Dugdale BSc ^105^, Richard Eccles MSc ^107^, Matthew Gemmell MSc ^107^, Dr Richard Gregory PhD ^107^, Dr Margaret Hughes PhD ^107^, Charlotte Nelson MSc ^107^, Dr Lucille Rainbow PhD ^107^, Dr Edith E Vamos PhD ^107^, Hermione J Webster BSc ^107^, Dr Mark Whitehead PhD ^107^, Claudia Wierzbicki BSc ^107^, Dr Adrienn Angyal PhD ^109^, Dr Luke R Green PhD ^109^, Dr Max Whiteley PhD ^109^, Emma Betteridge BSc ^116^, Dr Iraad F Bronner PhD ^116^, Ben W Farr BSc ^116^, Scott Goodwin MSc ^116^, Dr Stefanie V Lensing PhD ^116^, Shane A McCarthy ^116, 102^, Dr Michael A Quail PhD ^116^, Diana Rajan MSc ^116^, Dr Nicholas M Redshaw PhD ^116^, Carol Scott ^116^, Lesley Shirley MSc ^116^ and Scott AJ Thurston BSc ^116^

Software and analysis tools:

Dr Will Rowe PhD^43^, Amy Gaskin MSc ^74^, Dr Thanh Le-Viet PhD ^75^, James Bonfield BSc ^116^, Jennifier Liddle ^116^ and Andrew Whitwham BSc ^116^1 Barking, Havering and Redbridge University Hospitals NHS Trust, 2 Barts Health NHS Trust, 3 Belfast Health & Social Care Trust, 4 Betsi Cadwaladr University Health Board, 5 Big Data Institute, Nuffield Department of Medicine, University of Oxford, 6 Blackpool Teaching Hospitals NHS Foundation Trust, 7 Bournemouth University, 8 Cambridge Stem Cell Institute, University of Cambridge, 9 Cambridge University Hospitals NHS Foundation Trust, 10 Cardiff and Vale University Health Board, 11 Cardiff University, 12 Centre for Clinical Infection and Diagnostics Research, Department of Infectious Diseases, Guy’s and St Thomas’ NHS Foundation Trust, 13 Centre for Enzyme Innovation, University of Portsmouth, 14 Centre for Genomic Pathogen Surveillance, University of Oxford, 15 Clinical Microbiology Department, Queens Medical Centre, Nottingham University Hospitals NHS Trust, 16 Clinical Microbiology, University Hospitals of Leicester NHS Trust, 17 County Durham and Darlington NHS Foundation Trust, 18 Deep Seq, School of Life Sciences, Queens Medical Centre, University of Nottingham, 19 Department of Infectious Diseases and Microbiology, Cambridge University Hospitals NHS Foundation Trust, 20 Department of Medicine, University of Cambridge, 21 Department of Microbiology, Kettering General Hospital, 22 Department of Microbiology, South West London Pathology, 23 Department of Zoology, University of Oxford, 24 Division of Virology, Department of Pathology, University of Cambridge, 25 East Kent Hospitals University NHS Foundation Trust, 26 East Suffolk and North Essex NHS Foundation Trust, 27 East Sussex Healthcare NHS Trust, 28 Gateshead Health NHS Foundation Trust, 29 Great Ormond Street Hospital for Children NHS Foundation Trust, 30 Great Ormond Street Institute of Child Health (GOS ICH), University College London (UCL), 31 Guy’s and St. Thomas’ Biomedical Research Centre, 32 Guy’s and St. Thomas’ NHS Foundation Trust, 33 Hampshire Hospitals NHS Foundation Trust, 34 Health Services Laboratories, 35 Heartlands Hospital, Birmingham, 36 Hub for Biotechnology in the Built Environment, Northumbria University, 37 Hull University Teaching Hospitals NHS Trust, 38 Imperial College Healthcare NHS Trust, 39 Imperial College London, 40 Infection Care Group, St George’s University Hospitals NHS Foundation Trust, 41 Institute for Infection and Immunity, St George’s University of London, 42 Institute of Biodiversity, Animal Health & Comparative Medicine, 43 Institute of Microbiology and Infection, University of Birmingham, 44 Isle of Wight NHS Trust, 45 King’s College Hospital NHS Foundation Trust, 46 King’s College London, 47 Liverpool Clinical Laboratories, 48 Maidstone and Tunbridge Wells NHS Trust, 49 Manchester University NHS Foundation Trust, 50 Microbiology Department, Buckinghamshire Healthcare NHS Trust, 51 Microbiology, Royal Oldham Hospital, 52 MRC Biostatistics Unit, University of Cambridge, 53 MRC-University of Glasgow Centre for Virus Research, 54 Newcastle University, 55 NHS Greater Glasgow and Clyde, 56 NHS Lothian, 57 NIHR Health Protection Research Unit in HCAI and AMR, Imperial College London, 58 Norfolk and Norwich University Hospitals NHS Foundation Trust, 59 Norfolk County Council, 60 North Cumbria Integrated Care NHS Foundation Trust, 61 North Middlesex University Hospital NHS Trust, 62 North Tees and Hartlepool NHS Foundation Trust, 63 North West London Pathology, 64 Northumbria Healthcare NHS Foundation Trust, 65 Northumbria University, 66 NU-OMICS, Northumbria University, 67 Path Links, Northern Lincolnshire and Goole NHS Foundation Trust, 68 Portsmouth Hospitals University NHS Trust, 69 Public Health Agency, Northern Ireland, 70 Public Health England, 71 Public Health England, Cambridge, 72 Public Health England, Colindale, 73 Public Health Scotland, 74 Public Health Wales, 75 Quadram Institute Bioscience, 76 Queen Elizabeth Hospital, Birmingham, 77 Queen’s University Belfast, 78 Royal Brompton and Harefield Hospitals, 79 Royal Devon and Exeter NHS Foundation Trust, 80 Royal Free London NHS Foundation Trust, 81 School of Biological Sciences, University of Portsmouth, 82 School of Health Sciences, University of Southampton, 83 School of Medicine, University of Southampton, 84 School of Pharmacy & Biomedical Sciences, University of Portsmouth, 85 Sheffield Teaching Hospitals NHS Foundation Trust, 86 South Tees Hospitals NHS Foundation Trust, 87 Southwest Pathology Services, 88 Swansea University, 89 The Newcastle upon Tyne Hospitals NHS Foundation Trust, 90 The Queen Elizabeth Hospital King’s Lynn NHS Foundation Trust, 91 The Royal Marsden NHS Foundation Trust, 92 The Royal Wolverhampton NHS Trust, 93 Turnkey Laboratory, University of Birmingham, 94 University College London Division of Infection and Immunity, 95 University College London Hospital Advanced Pathogen Diagnostics Unit, 96 University College London Hospitals NHS Foundation Trust, 97 University Hospital Southampton NHS Foundation Trust, 98 University Hospitals Dorset NHS Foundation Trust, 99 University Hospitals Sussex NHS Foundation Trust, 100 University of Birmingham, 101 University of Brighton, 102 University of Cambridge, 103 University of East Anglia, 104 University of Edinburgh, 105 University of Exeter, 106 University of Kent, 107 University of Liverpool, 108 University of Oxford, 109 University of Sheffield, 110 University of Southampton, 111 University of St Andrews, 112 Viapath, Guy’s and St Thomas’ NHS Foundation Trust, and King’s College Hospital NHS Foundation Trust, 113 Virology, School of Life Sciences, Queens Medical Centre, University of Nottingham, 114 Watford General Hospital, 115 Wellcome Centre for Human Genetics, Nuffield Department of Medicine, University of Oxford, 116 Wellcome Sanger Institute, 117 West of Scotland Specialist Virology Centre, NHS Greater Glasgow and Clyde, 118 Whittington Health NHS Trust

